# Effect of Eight-Week Online Cognitive Training in Parkinson’s Disease: A Randomized Controlled Trial

**DOI:** 10.1101/2021.03.04.21252499

**Authors:** Tim D. van Balkom, Henk W. Berendse, Ysbrand D. van der Werf, Jos W.R. Twisk, Carel F.W. Peeters, Rob H. Hagen, Tanja Berk, Odile A. van den Heuvel, Chris Vriend

## Abstract

**Background:** Cognitive training (CT) has been proposed as a non-pharmacological treatment option for the frequent cognitive impairments occurring in PD.

**Objective:** Assess the efficacy of CT on cognitive function in PD.

**Methods:** In this double-blind, randomized controlled trial we enrolled 140 PD patients with significant subjective cognitive complaints. In eight weeks, participants underwent 24 sessions of computerized multi-domain CT or an active control condition for 45 minutes each (n=70 vs. n=70). The primary outcome was the accuracy on a computerized Tower of London task; secondary outcomes included effects on other neuropsychological outcomes and subjective cognitive complaints. Outcomes were assessed before and after training and at six-months follow-up, and were analyzed with multivariate mixed-model analyses.

**Results:** The intention-to-treat population consisted of 136 participants. Multivariate mixed-model analyses showed no group difference on the Tower of London accuracy corrected for baseline performance: B: −0.06, 95% CI: −0.27 to 0.15, p=0.562. Participants in the CT group were on average 0.30 SD (i.e., 1.5 seconds) faster on the Tower of London, difficulty load 4 (secondary outcome): 95% CI: −0.55 to −0.06, p=0.015. CT had similar positive effects on other processing speed-related executive function tasks, although these did not reach statistical significance. At follow-up, no group differences were present.

**Conclusions:** The results show tentative but consistent positive effects of CT on processing speed during executive functioning. Future studies should investigate booster sessions to increase durability, optimize training duration, and study different sub-groups of PD patients along the continuum of cognitive decline towards PD dementia.

## BACKGROUND

Cognitive impairment is highly prevalent in Parkinson’s disease (PD). At diagnosis already 25% of PD patients experience cognitive deficits in one or more domains[1] and the point prevalence of dementia in PD patients is 25-30%.[2] Estimates of the cumulative prevalence of PD dementia (PD-D) range from 46% after ten years of follow-up[3] to as high as 83% after twenty years.[4]

The available pharmacological treatments for cognitive impairment in PD have limited efficacy, focus on relieving symptoms but not on delaying decline, and can have negative side-effects.[2, 5] Cognitive training (CT) has been proposed as a promising alternative. CT may alleviate cognitive impairment and slow down cognitive decline by boosting neuroplasticity[6] and improving the efficiency of global and regional brain networks.[7]

Meta-analyses of previous CT studies in PD showed that CT has a small positive effect on global cognitive function.[8, 9] Larger effect sizes were reported for ‘frontal’ cognitive domains, including a moderate effect on executive function and small to moderate effects on working memory and mental processing speed (mean Hedges’ g ranging from 0·30-0·74). Two studies additionally showed long-term positive effects of CT, lasting up to 18 months after training,[10, 11] suggesting its potential in delaying cognitive decline. Nevertheless, the available evidence is based on small studies with methodological limitations; consequently, double-blind randomized controlled trials (RCT) using valid objective and subjective cognitive outcome measures are needed to provide more reliable evidence.[8, 12]

In this report we present the primary results of the COGnitive Training In Parkinson Study (COGTIPS).[13] We hypothesized, based on earlier research, that CT would predominantly improve executive function (primary outcome), but also other cognitive functions, i.e. working memory and processing speed. We also hypothesized that CT would decrease subjective cognitive complaints and would have long-term effects (i.e. at six-months follow-up).

## METHODS

### Trial design

COGTIPS is a large mono-center phase-3 double-blind RCT to assess superiority of eight-week computerized CT over an active control condition (AC). Participants were enrolled at the Amsterdam University Medical Centers (Amsterdam UMC), location VU University Medical Center. A detailed study protocol article was published before the end of the recruitment period and de-blinding, also including the results of a pilot feasibility study.[13] This trial was prospectively registered at ClinicalTrials.gov, identifier NCT02920632 (September 30, 2016) and the CONSORT checklist is provided as Supplementary Material.

### Participants

We enrolled 140 PD patients that were eligible for participation, with a) mild to moderately advanced idiopathic PD, diagnosed by a neurologist (Hoehn and Yahr stage < 4),[14] b) significant subjective cognitive complaints (Parkinson’s Disease Cognitive Functional Rating Scale (PD-CFRS) score > 3),[15] and c) home access to a computer or tablet with internet. Exclusion criteria were a) Montreal Cognitive Assessment (MoCA) score < 22,[16, 17] b) indications of current drug- or alcohol abuse (CAGE AID-interview score > 1),[18, 19] c) moderate or severe depressive symptoms (Beck Depression Inventory (BDI) score > 18),[20] d) impulse control disorder (positive screening by diagnostic criteria), e) psychotic symptoms (positive screening by the Schedule for Assessment of Positive Symptoms – PD),[21] except for benign hallucinations, or f) history of traumatic brain injury with loss of consciousness for > 15 min and/or posttraumatic amnesia > 1 h. All participants gave written informed consent and the study was approved by the Medical Ethical Committee of the VU University Medical Center.

### Randomization and blinding

Detailed information is provided in our design article.[13] Briefly, participants were randomized over an experimental CT condition and an AC in an 1:1 fashion. Randomization lists were generated using a random number sequence, stratified on education level. Blinding of participants was ensured by not providing any details to participants about the two conditions. Participants remained blind to their condition throughout the entire study and outcome assessors were blinded during all assessments. The training interventions were explained after participants completed the baseline assessment. Blinded study members (TB and trained research assistants) enrolled and assessed participants. Only non-blinded study members (CV and trained research assistants) had access to the allocation and randomization files.

### Procedures

In both conditions, participants followed an online home-based intervention on computer or tablet, that had a duration of eight weeks, three times a week for approximately 45 minutes (total duration: 1080 min). The CT consisted of 13 training games, that had an adaptive difficulty based on the individual participants’ performance, based on the ‘Braingymmer’ online CT platform (www.braingymmer.com, a product by Dezzel Media). The training aimed to improve ‘frontal’ cognitive functions. The games were not part of the pre- and post-intervention assessments. We corrected for non-specific cognitive engagement by using an AC that consisted of three games without difficulty adjustments (i.e., hangman, trivia questions and solitaire).

At baseline (T0), after training (T1, at approximately nine weeks) and at follow-up (T2, approximately six-months after training) patients underwent an extensive assessment that included neuropsychological tests, questionnaires and interviews (see below and the protocol article[13] for details).

### Outcomes

The primary outcome measure was the efficacy of CT, relative to the AC, measured with the percentage correct responses (i.e. accuracy) on a computerized self-paced version of the Tower of London (ToL) task.[22, 23] The ToL covers various executive functions including planning, inhibition, attention, and working memory and consists of 100 pseudo-randomized trials with varying difficulty, ranging from one-step to five-step solutions (task-load S1-S5). Participants were excluded from ToL data analysis if they showed poor understanding of the task, operationalized as a score < 75% on the basic one-step (S1) trials. We used reaction time on the ToL as secondary outcome. A detailed list of all assessment instruments is provided in the Supplementary Material (p 2).

Additional secondary outcomes were CT effects at T1 and T2 on subjective cognitive complaints and cognitive performance on latent cognitive factors (see Statistical methods). Subjective cognitive complaints were measured with the self-report and informant version of the PD-CFRS and the Cognitive Failures Questionnaire.[24] Latent cognitive factors were defined on the basis of an extensive neuropsychological test battery that mapped global cognitive function and performance on five cognitive domains, i.e., executive function, attention/speed of processing/working memory, episodic memory, language, and visuospatial/visuoconstructive function. Based on neuropsychological function compared with healthy norm groups (see Table S1 of the Supplementary Material) we classified cognitive function of patients as cognitively normal, cognitive deficits associated with level II Movement Disorder Society (MDS) criteria for PD-MCI[25] or cognitive features of probable PD-D.[26]

Exploratory outcomes included group differences in *individual* neuropsychological test scores, performance on the CT and AC games, and effects on psychiatric symptoms, including depression (BDI), anxiety (Parkinson Anxiety Scale),[27] apathy (Apathy Scale)[28] and impulse control disorders (Questionnaire for impulsive-compulsive disorders in Parkinson’s Disease-Rating Scale; QUIP-RS).[29]

No serious adverse events were expected from the interventions and assessments. We therefore only assessed adverse events related to impulse control disorders (including Internet addiction) for which PD patients are at increased risk.

### Statistical methods

The sample size calculation was based on a repeated-measures ANOVA corrected for a moderate correlation between pre- and post-intervention outcomes (*r*=0.6) and an effect size *f*=0.12 of CT on global cognitive function as reported in an earlier meta-analysis in PD patients.[8] The sample size needed to detect this effect at an α=0.05 and β=0.80 was n=112. To ensure adequate power – also at follow-up – and given a small expected drop-out, the desired sample size was set at n=140. We subsequently adopted a better analysis technique (i.e., multivariate mixed-model analysis) that could handle missing values and model multivariate effects to study group differences.

Analyses were performed on the intention-to-treat population (i.e., all correctly enrolled and randomized participants). We used the mean or median with standard deviations or range to present demographic and clinical variables. We used a multivariate linear mixed-model analysis to assess differences between groups on the primary outcome measure (ToL accuracy) with z-transformed mean accuracy scores on task-load S1-S5 (modelled together) at T1 as multivariate outcome. We modelled standardized mean accuracy scores of these measures at T0 as covariates, condition as independent variable and a random intercept at participant level to correct for correlation of the multiple variables within participants. In the intention-to-treat analysis, data of ten participants were missing due to failed assessment (n=4), no follow-up (n=2) or poor understanding (n=4) for the primary outcome measure. As this proportion was very small (< 5%), we performed the planned analysis without using multiple imputation. All analyses were repeated with adjustment for age, sex and years of education.

We performed similar multivariate mixed-models to assess differences between groups on secondary outcomes, i.e., differences on the ToL reaction time on S1-S5, subjective cognitive complaints, and latent cognitive factors, using standardized measures. To determine the latent cognitive factors, we performed a regularized maximum likelihood factor analysis (for a detailed description see Supplementary Material p 3) to compute individual scores on latent cognitive factors.[30] The effects of CT at six months follow-up were analyzed similarly to the above, but with time as an additional covariate in the mixed-model. Post-hoc, we modelled global cognitive function classification – i.e., cognitively normal (PD-NC), PD-MCI, or PD-D – as an additional covariate in tests that showed CT-induced change, to assess differential training effects between these subgroups.

We performed exploratory analyses of the CT effect on individual neuropsychological tests with univariate linear mixed-models using the performance at T1 as outcome, the performance at T0 as covariate and condition as independent variable. We analyzed the change in performance on the CT and AC games using multivariate mixed-model analyses and additionally assessed ceiling effects on the intervention by comparing six phases of training (session 1-4 compared with session 5-8, etc.; for a detailed description see Supplementary Material p 3). The effect of CT relative to the AC on psychiatric symptoms was analyzed using multivariate mixed-model analyses with standardized scores on the psychiatric symptom questionnaires at T1 as multivariate outcomes, the standardized T0 measurements as covariates and condition as independent variable.

We ran statistical analyses in SPSS version 26.0 (Armonk, NY, USA) and performed factor analysis using the FMradio package in R (version 3.5.3, Boston, MA, USA), using two-sided tests with statistical threshold of α < 0.05. We did not correct for multiple comparisons in our primary and secondary analyses as these involved four multivariate models for separate research questions. Exploratory analyses were separately corrected for multiple comparisons using a false discovery rate (q<0.05). During the trial the Clinical Research Bureau of Amsterdam UMC performed two data monitoring visits.

### Role of the funding organization

Two members of the Dutch Parkinson’s Disease Patient Association made a contribution to the design of the study. The funding bodies had no role in the collection, analysis, and interpretation of data, writing the manuscript, or the decision to submit the manuscript for publication. Dezzel Media B.V. did not sponsor this study, nor contributed to the design of the study, the analysis and interpretation of data, writing the manuscript or the decision to submit the manuscript for publication.

## RESULTS

### Participants

We enrolled 140 PD patients between September 15^th^ 2017 and May 23^rd^ 2019 with six-month follow-up assessments until January 29^th^ 2020. A flowchart is provided in Figure 1. Four participants were wrongfully enrolled and therefore excluded from the analyses. One-hundred-and-thirty-six (136) participants remained with mean age 62.9 years (SD=7.6) and 54 participants (39.7%) were female. Four participants (2.8%, two in either condition) dropped out during the intervention including one that underwent an exit measurement. One participant was lost to follow-up after the intervention (0.7%).

**Figure 1:**
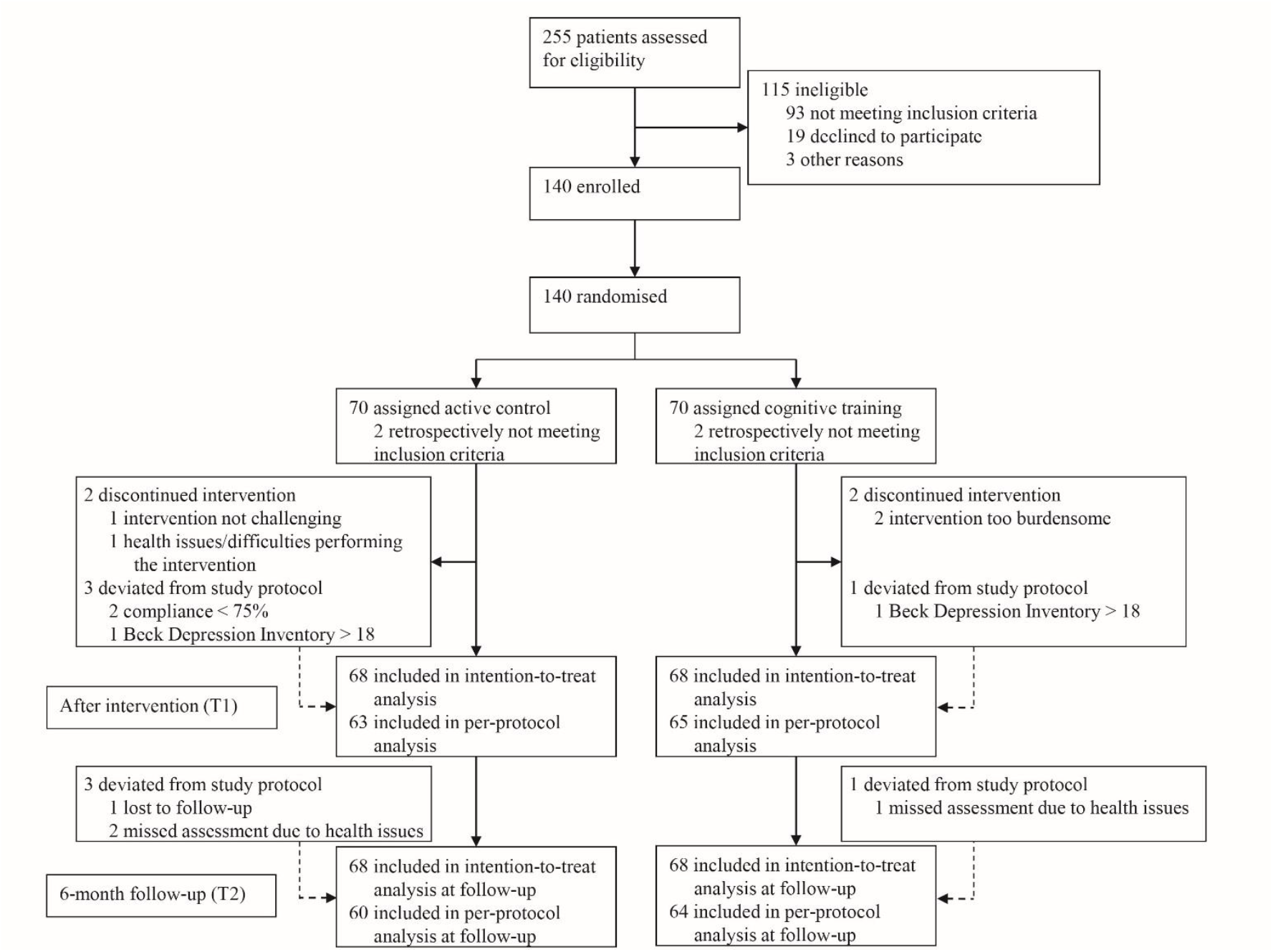
Flowchart of the enrollment procedure.

Demographic and clinical characteristics are provided in Table 1. There were small differences in sex distribution, education and baseline cognitive complaints between groups. The groups were similar on other demographic and clinical characteristics. Compared with healthy norm groups, the participants’ average cognitive performance was below average on attention and processing speed tasks, but normal for other cognitive domains (see Table S1 and Figure S1 in the Supplementary Material).

**Table 1:** Demographic and clinical characteristics of the intention-to-treat population.

|  | Active control (n=68) | Cognitive training (n=68) |
| --- | --- | --- |
| **Sex (N (%))** |  |  |
| Male | 47 (69%) | 35 (51%) |
| Female | 21 (31%) | 33 (49%) |
| **Age (years)** | 62.9 (7.0) | 62.9 (8.1) |
| **Education (years)** | 16.7 (4.4) | 15.5 (3.3) |
| **Education classification (N (%))<sup>a</sup>** |  |  |
| 3 | 1 (1.5%) | 1 (1.5%) |
| 4 | 4 (5.9%) | 4 (5.9%) |
| 5 | 16 (23.5%) | 17 (25.0%) |
| 6 | 26 (38.2%) | 29 (42.6%) |
| 7 | 21 (30.9%) | 17 (25.0%) |
| **Disease duration (years, median [range])** | 5 [1-26] | 5 [0-22] |
| **UPDRS-III** | 21.0 (9.5) | 20.2 (8.3) |
| **Hoehn & Yahr stage (N (%))** |  |  |
| 1 | 5 (7.4%) | 4 (5.9%) |
| 1.5 | 2 (2.9%) | 7 (10.3%) |
| 2 | 34 (50.0%) | 28 (41.2%) |
| 2.5 | 18 (26.5%) | 18 (26.5%) |
| 3 | 9 (13.2%) | 11 (16.2%) |
| **LEDD (median [range])** | 650 [0-2100] | 737 [0-1665] |
| **Medication change during study (N (%))** | 15 | 11 |
| **LEDD T1 (median [range], N=132)** | 710 [0-1981] | 762 [0-1530] |
| **MoCA** | 25.9 (2.3) | 26.3 (2.0) |
| **Global cognitive function classification (N (%))** |  |  |
| Normal cognition | 13 (19.1%) | 15 (22.1%) |
| Single-domain MCI | 7 (10.3%) | 9 (13.2%) |
| Multi-domain MCI | 35 (51.5%) | 34 (50.0%) |
| PD dementia | 13 (19.1%) | 10 (14.7%) |
| **BDI** | 7.87 (4.1) | 8.21 (4.0) |
| **QUIP-RS (N=125)** | 19.2 (12.7) | 15.8 (12.8) |
| **PAS (N=135)** | 10.5 (6.8) | 10.3 (6.6) |
| **AS (N=135)** | 13.4 (4.5) | 13.2 (4.5) |
| **Credibility-Expectancy (N=135)** | 32.7 (7.6) | 33.9 (6.0) |
| **PD-CFRS (median [range])** | 9.0 [3.3-22] | 7.0 [3.3-19] |
| **Compliance (%; median [range])** | 100 [25-100] | 100 [39-100] |
| **T0-to-T1 interval (days)** | 64.3 (6.5) | 63.6 (4.8) |
| **T0-to-T2 interval (days)** | 253 (14) | 250 (10) |

Data are mean (SD) unless otherwise specified. <sup>a</sup>According to Verhage education classification.<sup>29</sup>
**Abbreviations:** AS = Apathy Scale; BDI = Beck Depression Inventory; PAS = Parkinson Anxiety Scale; PD-CFRS = Parkinson's Disease – Cognitive Functional Rating Scale; LEDD = Levodopa equivalent daily dosage; MCI = mild cognitive impairment; MoCA = Montreal Cognitive Assessment; QUIP-RS = Questionnaire for Impulsive-Compulsive Disorders in Parkinson's Disease – Rating Scale; UPDRS = Unified Parkinson's Disease Rating Scale.

### Primary outcome – Tower of London accuracy

Below we only report the results of the intention-to-treat analysis. The analyses in the per-protocol sample showed similar results and are reported in Table S2 and S3 in the Supplementary Material. In the intention-to-treat sample, there was no difference between groups on ToL accuracy after training across all task-loads S1-S5 adjusted for baseline performance: B[SE]: −0.06 [0.10], 95% CI: −0.27 to 0.15, p=0.562 (crude model), or adjusted for baseline performance, age, sex and education level: B[SE]: −0.07 [0.10], 95% CI: −0.28 to 0.14, p=0.229 (adjusted model; see Figure 2a). The groups also showed no significant differences on the individual ToL accuracy task-loads (see Table 2).

**Table 2:** Group differences from the multivariate linear mixed-model analyses on the primary and secondary outcome measures for the crude and adjusted analysis models.

|  | Baseline |  | T1 |  | Group difference (crude model) |  |  | Group difference (adjusted model)<sup>a</sup> |  |  |
| --- | --- | --- | --- | --- | --- | --- | --- | --- | --- | --- |
|  | Active control<br>M (SD) | Cognitive training<br>M (SD) | Active control<br>M (SD) | Cognitive training<br>M (SD) | B [SE] | 95% CI | P value | B [SE] | 95% CI | P value |
| **Primary outcome measure** |  |  |  |  |  |  |  |  |  |  |
| Overall ToL accuracy (%)<sup>b</sup> | 82.3 (7.9) | 81.2 (9.1) | 84.9 (8.2) | 84.0 (10.4) | -0.060 [0.104] | -0.266 to 0.145 | 0.562 | -0.070 [0.103] | -0.275 to 0.135 | 0.501 |
| S1 | 96.5 (5.1) | 96.0 (5.9) | 96.9 (4.9) | 96.2 (4.7) | -0.118 [0.162] | -0.435 to 0.200 | 0.466 | -0.127 [0.161] | -0.444 to 0.190 | 0.430 |
| S2 | 92.0 (8.7) | 90.6 (9.1) | 92.5 (8.5) | 90.6 (11.6) | -0.140 [0.162] | -0.458 to 0.177 | 0.385 | -0.151 [0.161] | -0.468 to 0.166 | 0.350 |
| S3 | 87.7 (10.9) | 86.8 (12.0) | 88.8 (9.1) | 89.0 (13.2) | 0.039 [0.162] | -0.278 to 0.357 | 0.808 | 0.030 [0.161] | -0.287 to 0.347 | 0.852 |
| S4 | 75.9 (13.5) | 75.0 (15.2) | 80.8 (14.1) | 78.7 (17.2) | -0.112 [0.162] | -0.429 to 0.206 | 0.490 | -0.121 [0.161] | -0.437 to 0.196 | 0.455 |
| S5 | 59.7 (20.4) | 57.6 (20.0) | 65.3 (21.4) | 65.3 (22.2) | 0.029 [0.162] | -0.289 to 0.346 | 0.859 | 0.019 [0.161] | -0.298 to 0.336 | 0.906 |
| **Secondary outcome measures** |  |  |  |  |  |  |  |  |  |  |
| Overall ToL reaction time (s)<sup>c,d</sup> | 12.7 (2.7) | 12.4 (3.2) | 12.1 (2.9) | 11.4 (2.9) | -0.116 [0.097] | -0.308 to 0.076 | 0.232 | -0.116 [0.097] | -0.308 to 0.076 | 0.232 |
| S1 | 6.4 (1.9) | 5.9 (1.8) | 5.9 (2.1) | 5.3 (1.6) | -0.198 [0.124] | -0.443 to 0.046 | 0.111 | -0.150 [0.124] | -0.394 to 0.094 | 0.227 |
| S2 | 8.3 (2.6) | 8.1 (2.9) | 7.4 (2.1) | 7.1 (2.4) | -0.108 [0.124] | -0.352 to 0.135 | 0.382 | -0.057 [0.124] | -0.301 to 0.186 | 0.644 |
| S3 | 11.0 (2.7) | 11.0 (3.5) | 10.6 (3.3) | 10.0 (3.5) | -0.157 [0.124] | -0.401 to 0.086 | 0.204 | -0.105 [0.124] | -0.348 to 0.138 | 0.397 |
| S4 | 15.6 (3.7) | 15.6 (4.8) | 15.2 (4.8) | 13.8 (4.7) | -0.304 [0.124] | -0.548 to -0.060 | 0.015 | -0.252 [0.124] | -0.495 to -0.009 | 0.042 |
| S5 | 22.0 (4.9) | 21.5 (4.8) | 21.5 (4.3) | 20.9 (4.5) | -0.067 [0.124] | -0.312 to 0.177 | 0.588 | -0.016 [0.124] | -0.260 to 0.228 | 0.897 |
| Overall subjective cognitive complaints<sup>c,e</sup> |  |  |  |  | -0.06 [0.11] | -0.28 to 0.15 | 0.560 | -0.03 [0.11]* | -0.25 to 0.20 | 0.821 |
| PD-CFRS | 9.7 (4.7) | 8.0 (4.0) | 8.0 (5.1) | 6.9 (4.3) | -0.06 [0.14] | -0.33 to 0.22 | 0.689 | -0.02 [0.14]* | -0.30 to 0.26 | 0.880 |
| PD-CFRS inf. | 5.6 (4.3) | 6.0 (5.0) | 5.7 (4.3) | 5.3 (3.6) | 0.00 [0.17] | -0.34 to 0.34 | 0.981 | 0.04 [0.17]* | -0.30 to 0.39 | 0.798 |
| CFQ | 38.5 (11.5) | 38.2 (10.6) | 38.0 (12.7) | 36.9 (10.5) | -0.11 [0.14] | -0.38 to 0.17 | 0.437 | -0.07 [0.14]* | -0.34 to 0.21 | 0.635 |
| Overall cognitive factors<sup>b</sup> |  |  |  |  | 0.024 [0.068] | -0.110 to 0.158 | 0.722 | 0.002 [0.070] | -0.135 to 0.138 | 0.983 |
| Factor 1 | -0.035<br>(0.993) | 0.100<br>(1.125) | -0.002<br>(1.194) | 0.076<br>(0.947) | -0.002 [0.152] | -0.301 to 0.298 | 0.992 | -0.023 [0.152] | -0.322 to 0.276 | 0.879 |
| Factor 2 | -0.005<br>(1.252) | 0.117<br>(1.204) | -0.075<br>(1.236) | 0.134<br>(1.072) | 0.137 [0.152] | -0.162 to 0.437 | 0.368 | 0.116 [0.152] | -0.183 to 0.415 | 0.448 |
| Factor 3 | 0.025<br>(0.935) | -0.025<br>(0.905) | 0.096<br>(0.734) | -0.048<br>(1.081) | -0.115 [0.152] | -0.414 to 0.185 | 0.452 | -0.138 [0.152] | -0.437 to 0.161 | 0.365 |
| Factor 4 | 0.054<br>(1.065) | -0.002<br>(0.917) | -0.068<br>(1.080) | 0.081<br>(1.016) | 0.182 [0.152] | -0.118 to 0.481 | 0.234 | 0.158 [0.152] | -0.141 to 0.457 | 0.299 |
| Factor 5 | 0.042<br>(1.096) | -0.069<br>(1.107) | 0.087<br>(1.050) | -0.061<br>(1.062) | -0.081 [0.152] | -0.381 to 0.218 | 0.594 | -0.105 [0.152] | -0.404 to 0.194 | 0.490 |

<sup>a</sup>Corrected for age, sex and education in years; <sup>b</sup>Positive estimates indicate effects in favor of CT; <sup>c</sup>Negative estimates indicate effects in favor of CT; <sup>d</sup>Reaction time of correct responses;<sup>e</sup>Model additionally corrected for credibility/expectancy questionnaire score.
Abbreviations: CFQ = Cognitive Failures Questionnaire; inf. = informant version; PD-CFRS = Parkinson's disease – Cognitive Functional Rating Scale; ToL = Tower of London.

**Figure 2:**
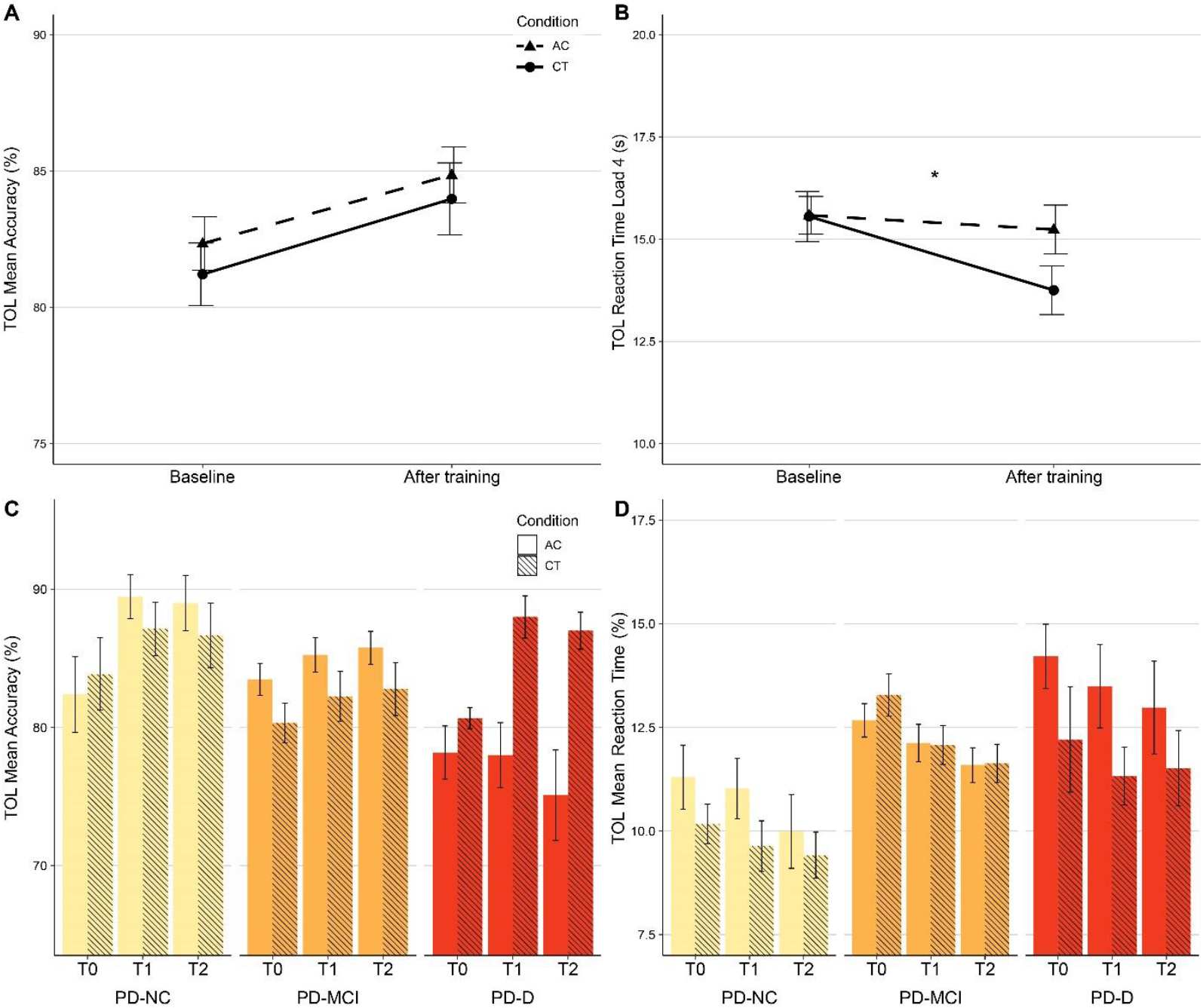
Intervention effect in the cognitive training (CT) and active control (AC) group. The upper panels show effects on the Tower of London (TOL) mean accuracy of S1-S5 **(A)** and reaction time **(B)**. *Indicates significant difference after training adjusted for baseline performance in the crude model. The lower panel shows results of the post-hoc analyses – the difference between intervention effects on the Tower of London (TOL) mean accuracy **(C)** and reaction time **(D)**, separated for participants with normal cognition (PD-NC), PD-mild cognitive impairment (PD-MCI) and PD dementia (PD-D). Data shown are observed means ± standard error.

### Secondary outcomes

Group differences and statistics on the secondary outcomes are depicted in Table 2. Multivariate analysis of the ToL reaction times across task loads S1-S5 (n=126) showed that the CT group was on average 0.12 standard deviation faster after training compared with the AC group. This improvement was not significant: B[SE]: −0.12 [0.10], 95% CI: −0.31 to 0.08, p=0.232 (crude model); B[SE]: −0.12 [0.10], 95% CI: −0.31 to 0.08, p=0.232 (adjusted model). The CT group showed a statistically significant improvement of 0.28 standard deviation (i.e., 1.5 seconds) on task-load S4 relative to the AC group: B[SE]: −0.30 [0.12], 95% CI: −0.55 to - 0.06, p=0.015 (crude model); B[SE]: −0.25 [0.12], 95% CI: −0.50 to 0.01, p=0.042 (adjusted model; see Figure 2b). Estimates of the other ToL task-loads indicated numerically similar positive effects of CT compared with the AC, although these effects were not significant. Subjective cognitive complaints (n=133) showed no between-group differences.

Factor analysis on all cognitive outcomes resulted in five latent factors, that represented episodic memory (F1), executive and visuospatial function (F2), planning ability (F3), processing speed (F4), and attention and working memory (F5). After the intervention, there were no between-group differences on any of the factors (Table 2). Further details are reported in the Supplementary Material (pp 9-10). We report exploratory univariate analyses of the individual neuropsychological test outcomes in Table S5 in the Supplementary Material, showing estimates that suggest improvement in the CT group on the Stroop Color-Word Test card II and III and improvement in the AC group on the Rey Complex Figure Test, that, however, did not survive correction for multiple comparisons.

At six-months follow-up, no between-group differences were observed on the ToL accuracy or on any of the secondary cognitive or exploratory psychiatric outcomes (see Table S12-14 in the Supplementary Material).

### Exploratory and post-hoc analyses

Exploratory analyses of improvement on the intervention games are provided in Table S6 in the Supplementary Material. Participants improved significantly on the training in both conditions and on all separate training games. Comparison of six sequential phases of the training showed that the CT group no longer improved after phase IV, while the AC group no longer improved after phase I (see Figure 3). There was a significant association between improvement on the CT games and pre-to-post training change on the Stroop Color-Word Test card II and card III (see Table S7 in the Supplementary Material). Analyses of the effect of CT on psychiatric symptoms are reported in the Supplementary Material (pp 14-16).

**Figure 3:**
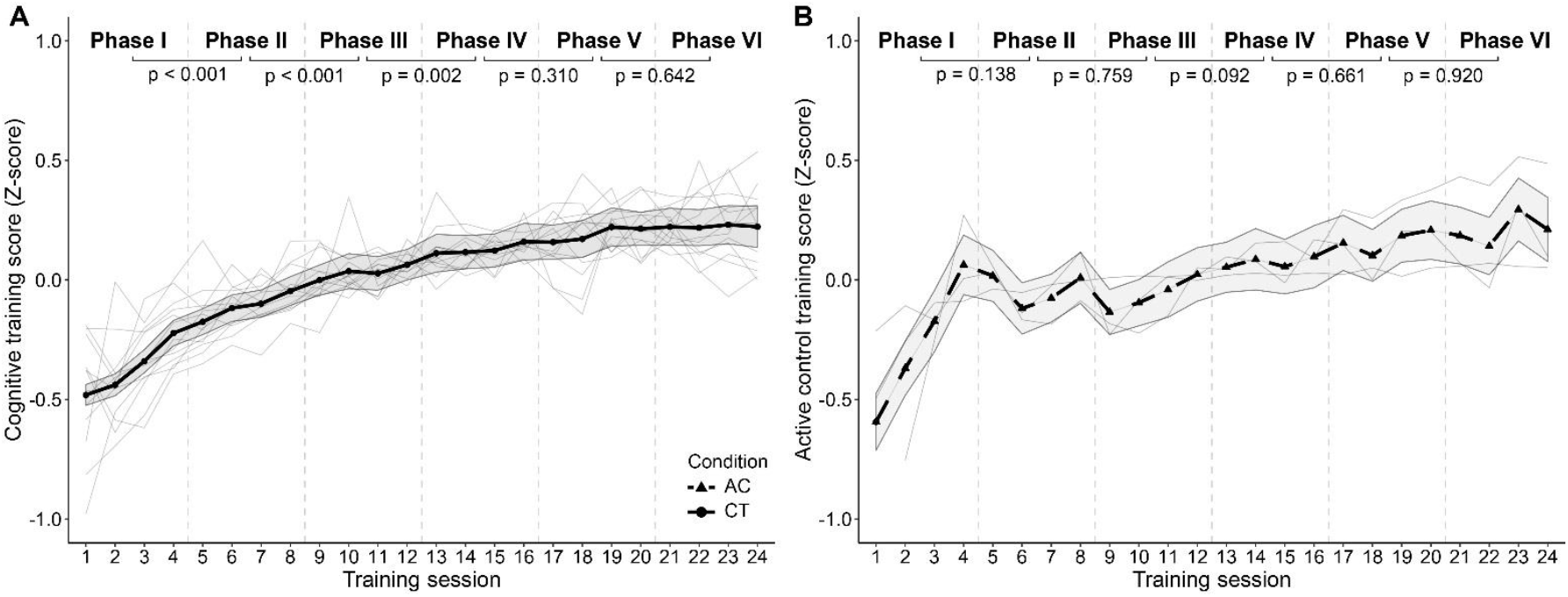
Improvement on cognitive training (CT; **A**) and active control (AC; **B**) training games. Mean game scores are shown per session with 95% CI, showing significant improvement with a ceiling effect. The lighter lines represent scores for individual games. The intervention period is divided in six phases of four sessions (marked with the vertical dotted lines). The *P* values provided in the figure are FDR-corrected and represent the difference between the two respective bins. Note that the Z-scores on the CT cannot be compared to the Z-scores on the active control as the CT games but not AC games were equipped with automatic difficulty adjustment, and the games of the two conditions were different.

Lastly, we performed post-hoc analyses of the differential effects of CT in separate cognitive diagnostic groups (PD-NC (n=28), PD-MCI (n=85) and PD-D (n=23)). The differential effects are illustrated in Figure 2 (ToL accuracy and reaction time) and Figure S3 (Stroop Color Word Test), and statistics are reported in Table S16 and Table S17 in the Supplementary Material. After training, CT effects were largest in the PD-D group. Interaction effects comparing the effect of CT between cognitive diagnostic groups showed significantly larger effects of CT for the PD-D group compared with the PD-NC and PD-MCI group for the ToL accuracy and Stroop Color-Word Test at T1. At T2, CT had a significantly larger positive effect on the ToL accuracy in the PD-D group compared with the other two diagnostic groups. Estimates of the intervention effect at T1 were all in favor of the CT for all diagnostic groups, except for ToL accuracy. The ToL accuracy showed positive estimates of the CT effect for the PD-D group but not for the PD-NC and PD-MCI group. At T2, there were no main or interaction effects for the ToL reaction time or Stroop Color Word Test at T2.

## DISCUSSION

In the COGTIPS double-blind RCT we assessed the efficacy of online home-based cognitive training (CT) in the largest sample of PD patients to date. Our results provide level I evidence that eight-week CT with BrainGymmer does not improve accuracy on a planning task (i.e., the primary outcome) in PD patients. On the secondary outcomes, CT showed consistent, but tentative, positive effects on measures of processing speed during executive functioning, that, however, only reached statistical significance for the ToL. The average improvement of speed was up to ten percent of the baseline performance. No effects were found in other cognitive domains or on subjective cognitive complaints. The observed positive effects of CT were no longer present at six-months follow-up.

Our study is in line with earlier aggregated findings from meta-analyses regarding the positive effects of CT on executive function and processing speed.[8, 9] Our multi-domain CT did not improve ToL accuracy, in contrast with an earlier, smaller study on CT in PD,[31] but the CT-related improvement on ToL reaction time presumably reflects both improved processing speed and planning function as this effect was driven by ToL items of higher difficulty.[32] The positive effects on the Stroop Color-Word Test are in line with earlier research[10] and our pilot feasibility study.[13] Stroop Color-Word Test improvement was related to improvement on the experimental intervention, likely due to the focus of the CT tasks on processing speed and executive functions. Surprisingly, the active control group performed better than the CT group after training on a visuoconstructive task, but these effects were very small (<1 point difference) and might be due to ceiling effects in the performance of this task.

Patients in both conditions reported minor subjective improvement after finishing the intervention, but without group differences. Interestingly, the positive effects of CT on processing speed measures did not positively affect scores on PD-specific questionnaires for subjective cognitive complaints, while mental slowness is a frequent early subjective complaint of PD patients.[33] Few studies that assessed the effects of CT on subjective complaints in PD found small improvements[10, 34] or null results.[31, 35, 36] Nevertheless, the sensitivity of the PD-CFRS in measuring treatment effects – as opposed to its sensitivity to measuring clinically relevant cognitive decline[15, 37] – has not yet been studied and our PD sample showed little variation in the total score at baseline. Limited transfer to ‘real-world’ cognitive function has previously been reported to be a shortcoming of CT and remains an important topic for future research.[38]

At six-months follow-up the group differences on the ToL reaction time and Stroop Color-Word Test that were present after training had levelled out. As yet, this does not support two earlier studies that assessed long-term CT effects.[10, 11] The 12- and 24-months follow-up assessments in our sample will provide more information. Our sample was largely comparable to PD samples in earlier CT studies, but participants had a relatively high mean years of education (16.1 years). Earlier studies reported a negative association between educational level and benefit from CT.[39, 40] A higher educational level generally reflects higher cognitive reserve and individuals with higher cognitive reserve reportedly retain intact cognitive function longer through compensatory neural mechanisms, but show cognitive decline at a faster rate.[41] If CT indeed positively impacts the underlying cognitive reserve, our one- and two-year follow-up assessments may show a delay in the onset or progression of cognitive decline in the CT group.

In both intervention groups, participants improved significantly on the intervention games (i.e., near transfer). Notably, the CT group improved despite increasing training difficulty. This supports the prerequisite that individuals with PD are trainable. Improvement on the CT reached a ceiling effect after approximately 20 sessions. There is limited literature on training dose effects; one study in PD patients suggested a “more-is-better” approach[42] while a meta-analysis in healthy adults showed that longer duration of training did not have larger effect sizes compared with shorter duration (i.e., longer/shorter than 20 hours).[43] The ceiling effect in our study may confirm the latter, but our data do not allow conclusions about the clinical relevance of training at the ceiling level. The null results at six-months follow-up indicate ‘booster’ training sessions may be necessary to prolong or strengthen the positive effects.

Our post-hoc tests suggested larger CT effects in PD-D patients compared with PD-NC and PD-MCI patients. Earlier studies did not differentiate CT effects between PD-NC, PD-MCI or PD-D patients. Our results were contrary to our expectation that an intervention in early-stage PD – when compensatory neural mechanisms may still be able to counteract progressive PD pathology – would be more efficacious.[13] The potential larger efficacy of CT in PD-D patients awaits replication in a larger sample as our PD-D sub-group was small. An explanation for the observed sub-group differences could be that cognitively normal PD participants in our sample performed at a ceiling level on cognitive tasks so that these tasks could not quantify improvement. Future analyses of the MRI scans in a subset of participants may be more sensitive and can show how compensatory neural mechanisms that are presumably present in the PD-NC and PD-MCI patients are affected by CT.

### Limitations and strengths

Limitations of our study were the lack of a measure for the level of cognitive activity and an additional waiting-list control group. Despite our eligibility criterion to exclude PD patients with severe cognitive impairments by using previously reported optimal diagnostic screening criteria for PD-D on the basis of the MoCA,[17] still a significant proportion of patients showed cognitive deficits associated with PD-D. Lastly, at baseline there were small, non-significant group differences in education level, sex distribution and subjective cognitive complaints, despite randomization and stratification. Although we adjusted for education and sex, this does not fully eliminate potential effects of inter-individual differences.

A major strength of our study was the sample size with an excellent intervention compliance and study protocol adherence. We used a prospectively registered, double-blind study, designed based on recommendations of earlier reviews.[8, 9] Participants underwent short- and long-term extensive neuropsychological assessments that adhered to the MDS Task Force guidelines,[25, 26] with additional motor and non-motor symptom assessment.

## Conclusions

This randomized, double-blind controlled trial in a large sample of PD patients showed that multi-domain CT did not improve accuracy on an executive function measure (primary outcome), but showed small but consistent effects on processing speed during executive functioning. The intervention was suitable for PD patients considering the high compliance, and participants showed large improvement on the training tasks. Ceiling effects after 16-20 sessions, combined with null results at six-months follow-up, may imply that for the clinical use of CT in PD patients shorter periods (i.e., ±20 sessions) of intensive CT combined with repetitive booster sessions should be studied.

## Supporting information

CONSORT

## Data Availability

The datasets generated, used and analyzed during the COGnitive Training In Parkinson Study are available from the corresponding author upon reasonable request.

## Authors’ Roles

1. Research project: A. Conception: TvB, HB, YvdW, RH, TB, OvdH, CV; B. Organization: TvB, HB, YvdW, OvdH, CV; C. Execution: TvB, OvdH, CV.
2. Statistical Analysis: A. Design: TvB, HB, YvdW, JT, CP, OvdH, CV; B. Execution: TvB, CV; C. Review and Critique: TvB, HB, YvdW, JT, CP, OvdH, CV.
3. Manuscript: A. Writing of the first draft: TvB, OvdH, CV; B. Review and Critique: TvB, HB, YvdW, JT, CP, RH, TB, OvdH, CV.

## Acknowledgments

We would like to thank drs. A.C.M. Kramer, A.L. Schrijer BSc., drs. A.M. Ticheler, A. van Weert BSc., drs. B.E. Olgers, D.N. van Deursen BSc., drs. E. Koedijk, drs. E.L. Vester, D.W. van Wylick BSc., drs. L. Drost, drs. F. Kooij, I. Ashour BSc., drs. I. Zijlstra, J. Breunese BSc., drs. J.S.R. Biesbroeck, drs. J.R.C. Verhaegh, K. Basant BSc., drs. M.J. Wagenmakers, mw. M.W. van der Wijk, drs. M.A. Laansma, M.M.A. Schyns BSc., drs. M. Rombouts, drs. M.G.M.S. Schokker, drs. J.P.A. van Dulm MD, and drs. S. Kasprzak for their invaluable work on the data collection.

We thank the COGTIPS user committee – dr. J.L.W. Bosboom MD, dr. G.J. Geurtsen, drs. W.J. Oudegeest MD, and drs. E. van der Rhee – for their input and suggestions in the execution and implementation of the trial.

We acknowledge the support from the Brain Foundation Netherlands.

Part of the participant recruitment was accomplished through Hersenonderzoek.nl, a Dutch online registry that facilitates participant recruitment for neuroscience studies (www.hersenonderzoek.nl). Hersenonderzoek.nl is funded by ZonMw-Memorabel (project no 73305095003), a project in the context of the Dutch Deltaplan Dementie, Gieskes-Strijbis Foundation, the Alzheimer’s Society in the Netherlands and Brain Foundation Netherlands. Participants were additionally recruited through ParkinsonNEXT, a Dutch online registry that aims to unite patients, researchers and clinicians wanting to contribute to research and innovation in Parkinson’s disease and Parkinsonism. ParkinsonNEXT produces information about ongoing studies and facilitates the recruitment of patients.

