## Supplementary Material for "Effect of Eight-Week Online Cognitive Training in Parkinson’s Disease: A Randomized Controlled Trial"

**Contents**


| | | |
|---|---|---|
| Supplement 1: | List of assessments instruments | p 2 |
| Supplement 2: | Supplementary methods | p 3 |
| Supplement 3: | Neuropsychological characteristics of the intention-to-treat sample | p 4 |
| Supplement 4: | Per-protocol sample description | p 7 |
| Supplement 5: | Analyses of the primary and secondary outcome measures in the per-protocol sample | p 8 |
| Supplement 6: | Results from the regularized redundancy-filtered factor analysis | p 9 |
| Supplement 7: | Exploratory univariate linear mixed-model analyses of neuropsychological test outcomes | p 11 |
| Supplement 8: | Improvement on the experimental and active control condition | p 12 |
| Supplement 9: | Association between CT and active control improvement and executive function change | p 13 |
| Supplement 10: | Analysis of the effect of CT on exploratory psychiatric outcomes | p 14 |
| Supplement 11: | Analyses of the neuropsychological and clinical measures at six-months follow-up in the intention-to-treat sample | p 17 |
| Supplement 12: | Analyses of the neuropsychological and clinical measures at six-months follow-up in the per-protocol sample | p 18 |
| Supplement 13: | Exploratory univariate linear mixed-model analyses of neuropsychological test outcomes at six-months follow-up | p 19 |
| Supplement 14: | Post-hoc analyses of differential CT effects in PD patients with normal cognition, mild cognitive impairment, or dementia | p 20 |
| References | | p 23 |

**1 List of assessments instruments**

| | |
|---|---|
| *Neuropsychological assessment* | |
| Global cognition | Montreal Cognitive Assessment[1] |
| | Pentagon copy from the Mini-Mental State Examination[2, 3] |
| Executive function | Computerized adaptation of the Tower of London[4] |
| | Controlled Oral Word Association Test ('letter fluency')[5] |
| | Stroop Color Word Test[6] – interference condition |
| Attention/speed of processing/working memory | Stroop Color Word Test[6] – word-naming and color-naming condition |
| | Wechsler Adult Intelligence Scale-III digit span[7] |
| Episodic memory | Rey Auditory Verbal Learning Test[8] |
| | Location Learning Test[9] |
| Language | Boston naming test[10] |
| | Category fluency[11] |
| Visuospatial/visuoconstructive function | Rey Complex Figure Test[12] |
| | Visual Form Discrimination Test[13] |
| *Questionnaires* | |
| Anxiety symptoms | Parkinson anxiety scale[14] |
| Apathy symptoms | Apathy scale[15] |
| Depressive symptoms | Beck depression inventory[16] |
| Impulse control disorder symptoms | Questionnaire for Impulsive-Compulsive Disorders in Parkinson's Disease – Rating Scale[17] |
| Subjective cognitive function | Parkinson's Disease – Cognitive Functional Rating Scale[18] |
| | Cognitive Failures Questionnaire[19] |
| Intervention expectation | Credibility/expectancy questionnaire[20] |
| *Demographic and other clinical characteristics* | |
| | Age |
| | Sex |
| Education level | Years of education |
| | Education level according to Verhage[21] |
| Disease onset | Age at disease onset |
| Motor symptom severity | Unified Parkinson's Disease Rating Scale – part III[22] |
| Disease stage | Hoehn and Yahr stage[23] |
| Medication use | Levodopa equivalent daily dose (LEDD) |

### 2 Supplementary methods

*Regularized redundancy-filtered factor analysis*

We performed a regularized maximum likelihood factor analysis on all cognitive outcomes at baseline.[24] We performed redundancy filtering at an (absolute value) threshold t=0.95 to account for item redundancy in the raw correlation matrix. Subsequently, a penalized maximum likelihood estimate of the filtered correlation matrix was obtained. The optimal value of the penalty parameter was determined by 5-fold cross-validation. We then assessed factorizability of the regularized correlation matrix and its individual variables using the Kaiser-Meyer-Olkin (KMO) measure and removed variables that modelled poorly, i.e. KMO < 0.9. We subsequently performed maximum likelihood factor analytic data compression on the resulting penalized correlation matrix and calculated the optimal factor solution using Guttman bounds. In the resulting factor model we assessed the proportion of explained variance. Lastly, we calculated squared multiple correlations between the observed variables and the latent common factors (which indicate indeterminacy of factor scores if < 0.9) and defined individual factor scores at T0, T1 and T2 using standardized Z-scores.

*Analysis of improvement on the intervention*

To analyze the improvement in performance on the CT and active control games, we first standardized training scores of the individual training games to Z-scores. We performed a multivariate mixed-model analysis per group (CT or active control) with the thirteen or three training games, respectively, as multivariate outcome, and session (ranging from 1-24) as independent variable. To assess potential ceiling effects, we divided the intervention period in six bins of four training sessions and added these bins as covariates to the model to assess the difference between the intervention stages (session 1-4 compared with session 5-8 and so on). We performed a post-hoc analysis to assess the association between improvement on training games and improvement on cognitive performance. We first calculated the linear slope coefficient of improvement during 24 training sessions on the 13 (CT condition) or three (active control condition) intervention games. We then added this coefficient as a covariate to mixed-model analyses of neuropsychological tests that showed change in the CT group.

### 3 Neuropsychological characteristics of the intention-to-treat sample

**Table S1:** Neuropsychological function based on healthy norms (intention-to-treat sample).

| Baseline | Total | Active control | Cognitive training | *P* value† |
|---|---|---|---|---|
| SCWT card I (N=136) | 35.25 (10.25) | 34.76 (11.49) | 35.74 (8.91) | 0.583 |
| SCWT card II (N=136) | 39.49 (10.33) | 39.32 (10.14) | 39.66 (10.59) | 0.849 |
| SCWT card III (N=136) | 44.73 (10.13) | 45.62 (10.22) | 43.84 (10.04) | 0.308 |
| SCWT interference score (N=136) | 53.00 (8.64) | 54.19 (8.66) | 51.81 (8.51) | 0.108 |
| Letter fluency (N=136) | 48.03 (11.26) | 48.43 (11.95) | 47.63 (10.59) | 0.116 |
| Category fluency (N=135) | 47.47 (9.77) | 48.78 (9.82) | 46.13 (9.61) | 0.116 |
| RCFT (N=136) | 46.27 (15.79) | 44.86 (15.70) | 47.68 (15.87) | 0.300 |
| RAVLT direct recall (N=136) | 43.60 (12.02) | 43.96 (12.70) | 43.25 (11.38) | 0.733 |
| RAVLT delayed recall (N=136) | 44.74 (12.07) | 44.94 (12.07) | 44.54 (12.16) | 0.849 |
| RAVLT delayed recall corrected for direct recall score (N=136) | 48.55 (11.76) | 48.38 (12.12) | 48.72 (11.48) | 0.868 |
| Digit span forward score (N=136) | 34.28 (6.91) | 34.37 (7.24) | 34.19 (6.62) | 0.882 |
| Digit span backward score (N=136) | 41.71 (7.21) | 42.40 (7.42) | 41.01 (6.98) | 0.265 |
| LLT learning curve (N=135)[25] | 46.13 (9.88) | 45.16 (9.71) | 47.10 (10.04) | 0.255 |
| LLT direct recall (N=135)[25] | 48.27 (8.42) | 47.84 (7.12) | 48.72 (9.59) | 0.546 |
| LLT delayed recall (N=134)[25] | 53.91 (8.08) | 53.89 (8.73) | 53.93 (7.46) | 0.982 |
| BNT (N=136)[26] | 52.61 (9.14) | 52.66 (8.56) | 52.56 (9.76) | 0.948 |
| BVFDT (N=136)[27] | 49.00 (9.79) | 48.34 (11.29) | 49.66 (8.04) | 0.432 |

*T-test. Norm scores are "T-scores" (M: 50, SD: 10) based on norms provided in Schmand and colleagues, unless otherwise specified.[28]
*Abbreviations:* BNT – Boston Naming Test; BVFDT – Benton Visual Form Discrimination Test; LLT – Location Learning Test; RAVLT – Rey Auditory Verbal Learning Test; RCFT – Rey Complex Figure Test; SCWT – Stroop Color Word Test.

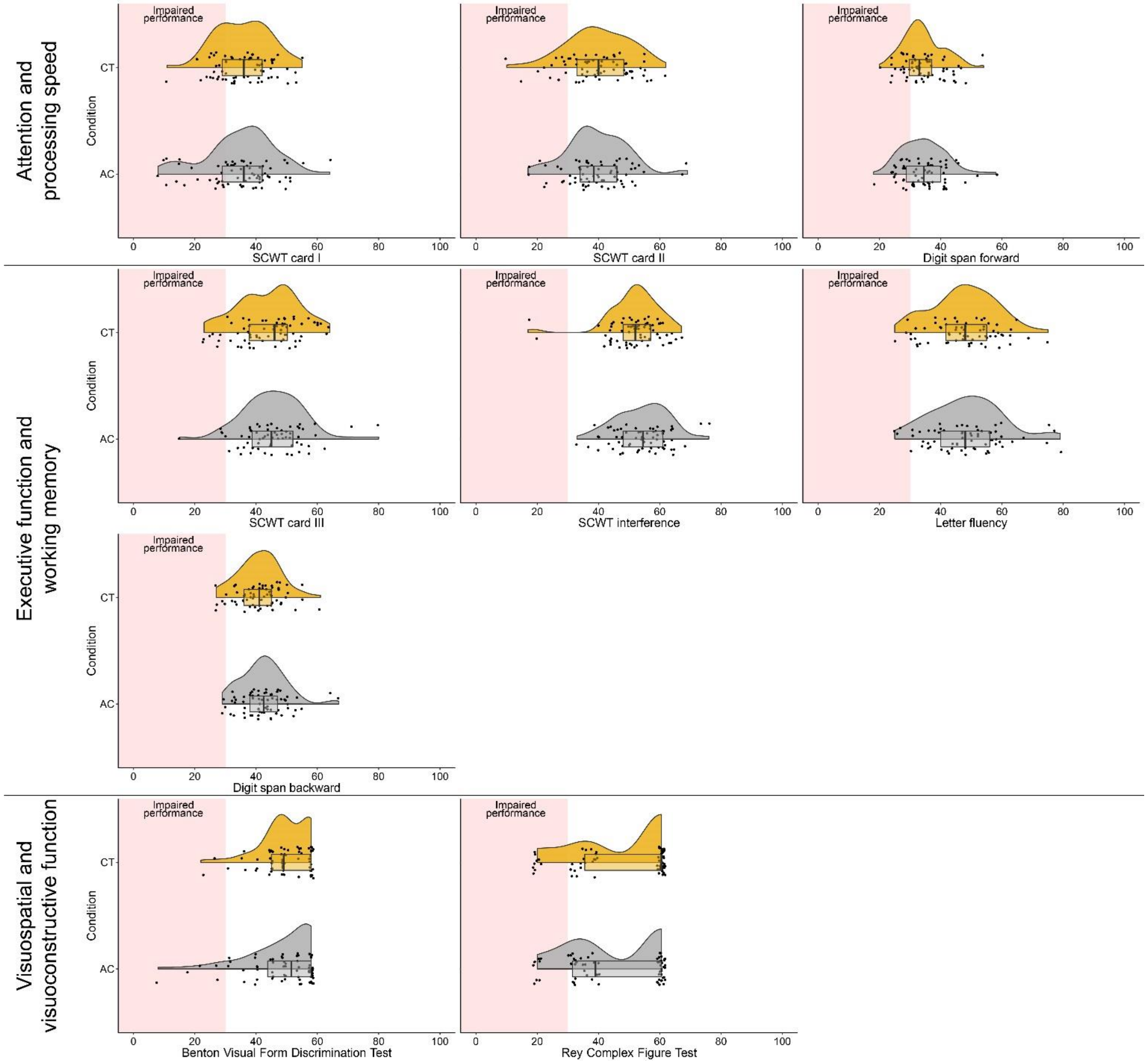


**Figure S1a:** Raincloud plots of neuropsychological test scores corrected for age, sex and/or education, using the appropriate norms. Test scores are provided as “T-scores” (M: 50, SD: 10). T-scores < 30 (i.e. > 2 SD below average) indicate impaired performance. *Abbreviations:* SCWT – Stroop Color Word Test.

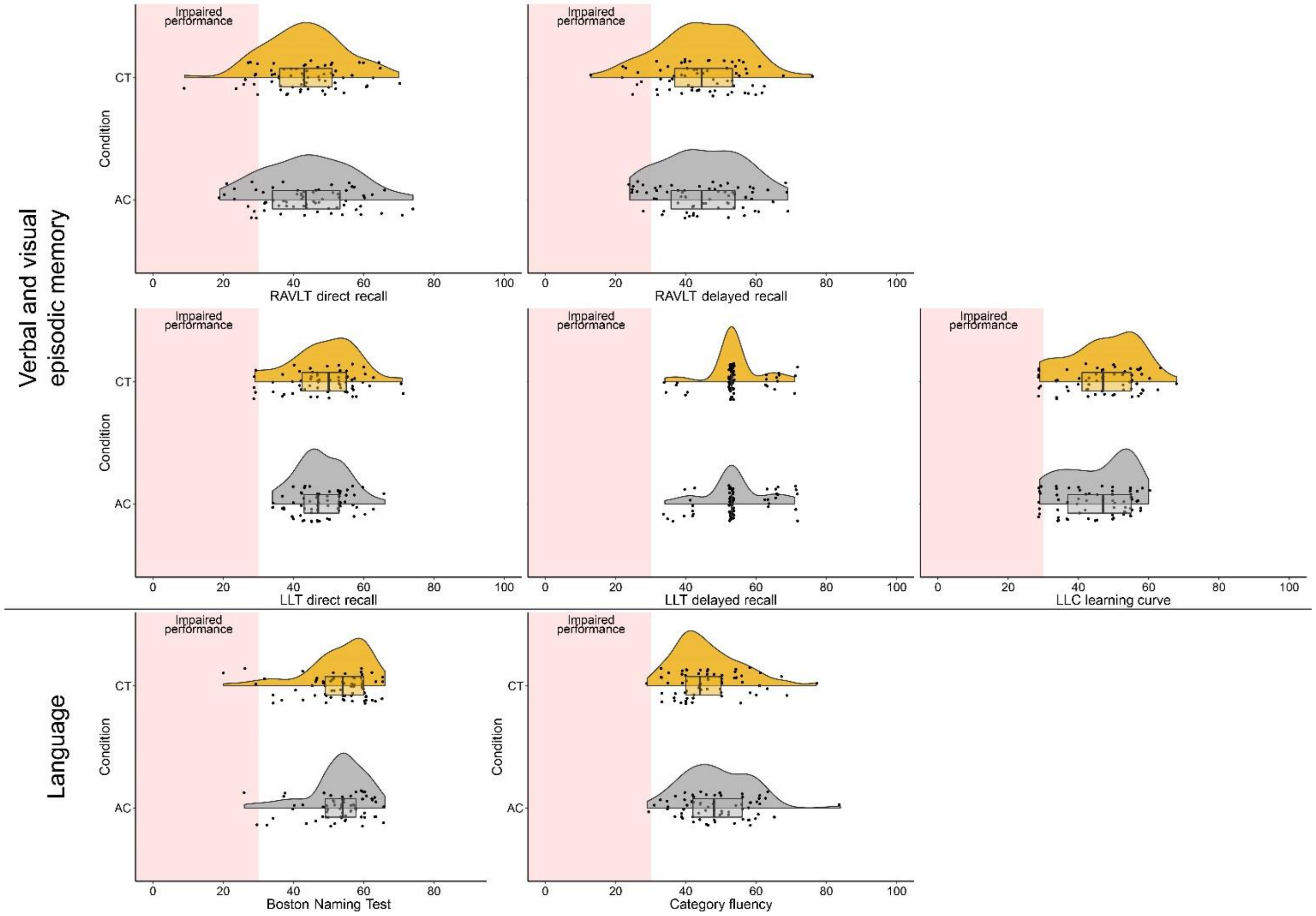


**Figure S1b:** Raincloud plots of neuropsychological test scores corrected for age, sex and/or education, using the appropriate norms. Test scores are provided as “T-scores” (M: 50, SD: 10). T-scores < 30 (i.e. > 2 SD below average) indicate impaired performance. *Abbreviations:* LLT – Location Learning Test; RAVLT – Rey Auditory Verbal Learning Test;

### 4 Per-protocol sample description

**Table S2:** Demographic and clinical characteristics of the per-protocol sample.

| | **Active control (n=63)** | **Cognitive training (n=65)** |
|---|---|---|
| **Sex (N (%))** | | |
| **Male** | 43 (68%) | 34 (52%) |
| **Female** | 20 (32%) | 31 (48%) |
| **Age (years)** | 63.0 (6.9) | 62.9 (8.1) |
| **Education (years)** | 16.9 (4.4) | 15.4 (3.2) |
| **Education classification (N (%))**† | | |
| **3** | 1 (1.6%) | 1 (1.5%) |
| **4** | 2 (3.2%) | 4 (6.2%) |
| **5** | 15 (23.8%) | 17 (26.2%) |
| **6** | 25 (39.7%) | 27 (41.5%) |
| **7** | 20 (31.7%) | 16 (24.6%) |
| **Disease length (years, median [range])** | 5 [1-26] | 5 [0-22] |
| **UPDRS-III** | 21.3 (9.0) | 20.6 (8.2) |
| **Hoehn & Yahr stage (N (%))** | | |
| **1** | 3 (4.8%) | 3 (4.6%) |
| **1.5** | 2 (3.2%) | 7 (10.8%) |
| **2** | 33 (52.4%) | 27 (41.5%) |
| **2.5** | 17 (27.0%) | 17 (26.2%) |
| **3** | 8 (12.7%) | 11 (16.9%) |
| **LEDD (median [range])** | 600 [0-2081] | 749 [0-1665] |
| **Medication change (N (%))** | 15 (24%) | 11 (17%) |
| **LEDD T1 (median [range])** | 700 [0-1981] | 750 [0-1530] |
| **MoCA** | 26.1 (2.3) | 26.4 (1.9) |
| **Global cognitive function classification (N (%)** | | |
| **Normal cognition** | 13 (20.6%) | 15 (23.1%) |
| **Single-domain MCI** | 7 (11.1%) | 9 (13.8%) |
| **Multi-domain MCI** | 31 (49.2%) | 32 (49.2%) |
| **PD dementia** | 12 (19.0%) | 9 (13.8%) |
| **BDI** | 7.9 (3.8) | 8.1 (3.9) |
| **QUIP-RS (N = 118)** | 18.8 (12.9) | 16.1 (12.9) |
| **PAS (N = 127)** | 10.5 (6.8) | 10.3 (6.8) |
| **AS (N = 127)** | 13.6 (4.4) | 13.3 (4.5) |
| **Credibility-Expectancy (N = 127)** | 32.7 (7.3) | 33.9 (6.1) |
| **PD-CFRS (median [range])** | 9.0 [3.3-22] | 7.2 [4-19] |
| **Compliance (%, median [range])** | 100 [80-100] | 100 [76-100] |
| **T0-to-T1 interval (days)** | 64.5 (6.6) | 63.7 (4.8) |
| **T0-to-T2 interval (days)** | 252 (14) | 250 (10) |

Data are mean (SD) unless otherwise specified. †According to Verhage education classification.[21] *Abbreviations*: AS = Apathy Scale; BDI = Beck Depression Inventory; PAS = Parkinson Anxiety Scale; PD-CFRS = Parkinson's Disease – Cognitive Functional Rating Scale; LEDD = Levodopa equivalent daily dosage; MCI = mild cognitive impairment; MoCA = Montreal Cognitive Assessment; QUIP-RS = Questionnaire for Impulsive-Compulsive Disorders in Parkinson's Disease – Rating Scale; UPDRS = Unified Parkinson's Disease Rating Scale.

### 5 Analyses of the primary and secondary outcome measures in the per-protocol sample

**Table S3:** Group differences from the multivariate linear mixed-model analyses on the primary and secondary outcome measures for the crude and adjusted analysis models in the per-protocol sample.

| | | **Baseline** | | **T1** | | **Group difference (crude model)** | | | **Group difference (adjusted model)*** | | |
|---|---|---|---|---|---|---|---|---|---|---|---|
| | | Active control M (SD) | Cognitive training M (SD) | Active control M (SD) | Cognitive training M (SD) | B [SE] | 95% CI | *P* value | B [SE] | 95% CI | *P* value |
| **Primary outcome measure** | | | | | | | | | | | |
| Overall ToL accuracy (%, N=121) | | 82.5 (7.9) | 81.0 (9.1) | 85.4 (8.0) | 83.9 (10.5) | -0.097 [0.104] | -0.303 to 0.108 | 0.351 | -0.112 [0.103] | -0.316 to 0.093 | 0.281 |
| Sub-score (%): | S1 | 96.3 (5.2) | 96.1 (5.9) | 97.1 (4.7) | 96.5 (4.4) | -0.121 [0.160] | -0.435 to 0.194 | 0.452 | -0.134 [0.159] | -0.447 to 0.179 | 0.401 |
| | S2 | 91.9 (8.8) | 90.8 (9.0) | 93.1 (8.1) | 91.0 (11.1) | -0.172 [0.160] | -0.487 to 0.142 | 0.282 | -0.187 [0.159] | -0.500 to 0.126 | 0.242 |
| | S3 | 88.0 (11.1) | 86.5 (12.1) | 89.4 (8.6) | 88.8 (13.3) | -0.025 [0.160] | -0.340 to 0.289 | 0.875 | -0.040 [0.159] | -0.353 to 0.273 | 0.802 |
| | S4 | 76.2 (13.6) | 74.9 (15.3) | 81.6 (13.9) | 78.3 (17.2) | -0.177 [0.160] | -0.492 to 0.137 | 0.268 | -0.192 [0.159] | -0.505 to 0.122 | 0.230 |
| | S5 | 60.2 (20.4) | 56.8 (19.7) | 65.6 (20.6) | 64.8 (22.3) | 0.010 [0.160] | -0.305 to 0.325 | 0.950 | -0.005 [0.160] | -0.319 to 0.308 | 0.973 |
| Secondary outcome measures | | | | | | | | | | | |
| Overall ToL reaction time (s, N=121)# | | 12.6 (2.8) | 12.5 (3.2) | 12.1 (2.9) | 11.4 (2.9) | -0.162 [0.096] | -0.353 to 0.028 | 0.094 | -0.114 [0.097] | -0.305 to 0.077 | 0.240 |
| Sub-score (s): | S1 | 6.3 (1.9) | 5.9 (1.7) | 5.9 (2.1) | 5.3 (1.6) | -0.186 [0.124] | -0.430 to 0.058 | 0.135 | -0.140 [0.124] | -0.385 to 0.104 | 0.259 |
| | S2 | 8.3 (2.6) | 8.2 (2.9) | 7.4 (2.1) | 7.1 (2.4) | -0.099 [0.124] | -0.342 to 0.145 | 0.427 | -0.050 [0.124] | -0.295 to 0.194 | 0.684 |
| | S3 | 10.9 (2.7) | 11.1 (3.5) | 10.6 (3.3) | 10.1 (3.6) | -0.174 [0.124] | -0.418 to 0.070 | 0.161 | -0.124 [0.124] | -0.368 to 0.120 | 0.317 |
| | S4 | 15.6 (3.8) | 15.6 (4.8) | 15.2 (4.7) | 13.8 (4.7) | **-0.294 [0.124]** | **-0.538 to -0.050** | **0.018** | **-0.245 [0.124]** | **-0.489 to -0.001** | **0.049** |
| | S5 | 22.0 (4.9) | 21.6 (4.8) | 21.5 (4.2) | 21.0 (4.5) | -0.058 [0.124] | -0.302 to 0.187 | 0.643 | -0.010 [0.124] | -0.254 to 0.235 | 0.939 |
| Overall subjective cognitive complaints (N=128) | | | | | | -0.05 [0.11] | -0.27 to 0.17 | 0.659 | -0.01 [0.11]† | -0.24 to 0.21 | 0.905 |
| Domain: | PD-CFRS | 9.5 (4.6) | 8.4 (3.9) | 8.1 (5.2) | 7.1 (4.2) | -0.08 [0.14] | -0.35 to 0.20 | 0.583 | -0.05 [0.14]† | -0.33 to 0.24 | 0.751 |
| | PD-CFRS inf. | 5.5 (4.1) | 6.2 (5.2) | 5.5 (4.1) | 5.4 (3.6) | 0.04 [0.17] | -0.31 to 0.38 | 0.841 | 0.07 [0.18]† | -0.28 to 0.42 | 0.686 |
| | CFQ | 38.8 (11.3) | 38.7 (10.7) | 38.1 (12.1) | 37.2 (10.3) | -0.07 [0.14] | -0.34 to 0.21 | 0.639 | -0.03 [0.14]† | -0.31 to 0.25 | 0.848 |
| Overall cognitive factors (N=117) | | | | | | 0.015 [0.070] | -0.123 to 0.152 | 0.832 | -0.004 [0.071] | -0.145 to 0.136 | 0.950 |
| Factors: | Factor 1 | -0.019 (0.975) | 0.088 (1.142) | 0.011 (1.198) | 0.073 (0.964) | 0.000 [0.156] | -0.306 to 0.306 | 0.999 | -0.018 [0.156] | -0.325 to 0.288 | 0.906 |
| | Factor 2 | 0.033 (1.267) | 0.127 (1.169) | -0.099 (1.253) | 0.146 (1.050) | 0.191 [0.156] | -0.116 to 0.497 | 0.222 | 0.172 [0.156] | -0.134 to 0.478 | 0.270 |
| | Factor 3 | 0.017 (0.954) | -0.028 (0.919) | 0.148 (0.706) | -0.049 (1.100) | -0.170 [0.156] | -0.476 to 0.136 | 0.276 | -0.190 [0.156] | -0.496 to 0.116 | 0.224 |
| | Factor 4 | 0.046 (1.070) | -0.016 (0.927) | -0.052 (1.079) | 0.063 (1.029) | 0.151 [0.156] | -0.155 to 0.457 | 0.333 | 0.131 [0.156] | -0.175 to 0.437 | 0.401 |
| | Factor 5 | 0.024 (1.093) | -0.101 (1.113) | 0.063 (1.047) | -0.107 (1.045) | -0.097 [0.156] | -0.403 to 0.209 | 0.535 | -0.117 [0.156] | -0.424 to 0.189 | 0.452 |

Significant differences are marked in bold. *Corrected for age, sex and education in years; #Reaction time of correct responses; †Model additionally corrected for credibility/expectancy questionnaire score.
Abbreviations: CFQ = Cognitive Failures Questionnaire; inf. = informant version; PD-CFRS = Parkinson's disease – Cognitive Functional Rating Scale; ToL = Tower of London.

**6 Results from the regularized redundancy-filtered factor analysis**

No outcomes were eliminated from the analysis due to redundant information. One outcome (i.e. Pentagon copy) was removed from analysis due to extremely low variance (96% scored 2/2, 3% scored 1/2) and the final Kaiser-Meyer-Olkin index was 0.960 which is indicative of great factorability.[29] Estimated image dimensions using Gutmann bounds showed an optimal factor solution of five factors with a cumulative explained variance of 80%. This factor solution is illustrated in Figure S1 and individual factor statistics are shown in Table S1. Squared multiple correlations between the observed features and five latent factors were all larger than 0.9, indicative of good determinacy of factor scores.

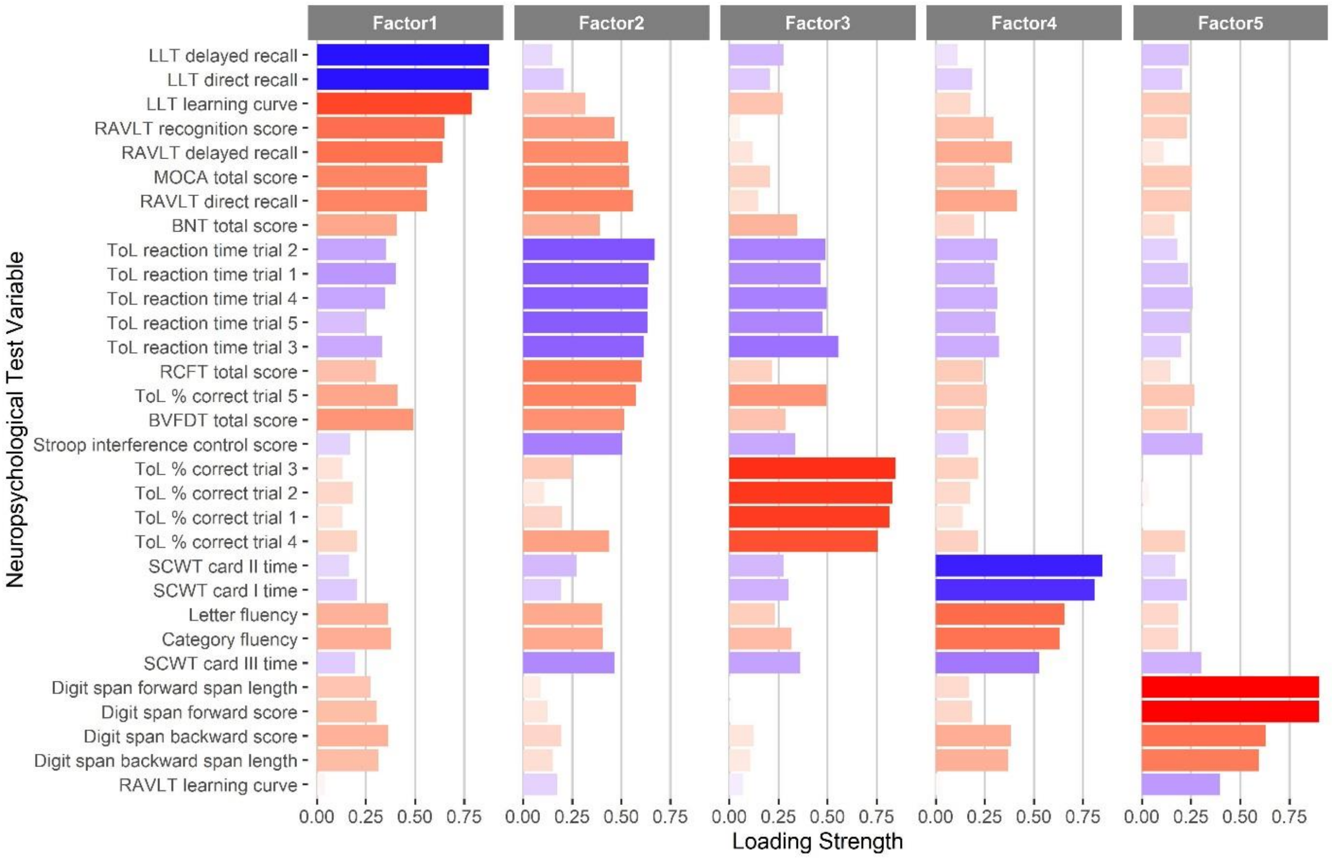


**Figure S2:** Bar graph of the five-factor solution. Length of the bars indicate the load strength on the specific factor. Color of the bars indicate a negative (i.e. blue) or positive (i.e. red) load.

**Table S4:** Five-factor solution with factor statistics.

| | **Episodic memory** | **Executive and visuospatial function** | **Planning ability** | **Processing speed** | **Attention and working memory** |
|---|---|---|---|---|---|
| **Sum of squares** | 5.78 | 5.75 | 5.24 | 4.28 | 3.70 |
| **Proportion explained variance** | 0.19 | 0.19 | 0.17 | 0.14 | 0.12 |
| **Squared multiple correlations** | 0.95 | 0.90 | 0.92 | 0.93 | 0.96 |
| **Neuropsychological test outcome** | | | | | |
| LLT delayed recall | **-0.876** | -0.148 | -0.275 | -0.113 | -0.237 |
| LLT direct recall | **-0.874** | -0.206 | -0.207 | -0.186 | -0.203 |
| LLT learning curve | **0.789** | 0.319 | 0.273 | 0.179 | 0.246 |
| RAVLT recognition | **0.651** | **0.464** | 0.052 | 0.294 | 0.230 |
| RAVLT delayed recall | **0.641** | **0.534** | 0.120 | 0.387 | 0.110 |
| MOCA total score | **0.560** | **0.538** | 0.206 | 0.299 | 0.256 |
| RAVLT direct recall | **0.560** | **0.562** | 0.146 | **0.414** | 0.250 |
| BNT total score | **0.406** | 0.391 | 0.346 | 0.196 | 0.166 |
| ToL reaction time trial 2 | -0.351 | **-0.670** | **-0.490** | -0.314 | -0.181 |
| ToL reaction time trial 1 | **-0.404** | **-0.640** | **-0.465** | -0.299 | -0.236 |
| ToL reaction time trial 4 | -0.347 | **-0.636** | **-0.496** | -0.313 | -0.259 |
| ToL reaction time trial 5 | -0.245 | **-0.634** | **-0.475** | -0.304 | -0.241 |
| ToL reaction time trial 3 | -0.332 | **-0.617** | **-0.555** | -0.322 | -0.200 |
| RCFT total score | 0.302 | **0.604** | 0.218 | 0.241 | 0.144 |
| ToL % correct trial 5 | **0.410** | **0.573** | **0.494** | 0.262 | 0.266 |
| BVFDT total score | **0.492** | **0.514** | 0.285 | 0.250 | 0.231 |
| SCWT interference control | -0.170 | **-0.504** | -0.334 | -0.165 | -0.308 |
| ToL % correct trial 3 | 0.131 | 0.249 | **0.847** | 0.217 | -0.001 |
| ToL % correct trial 2 | 0.183 | 0.108 | **0.829** | 0.175 | 0.037 |
| ToL % correct trial 1 | 0.130 | 0.196 | **0.816** | 0.139 | -0.013 |
| ToL % correct trial 4 | 0.205 | **0.438** | **0.754** | 0.215 | 0.220 |
| SCWT card II time | -0.163 | -0.272 | -0.275 | **-0.848** | -0.169 |
| SCWT card I time | -0.205 | -0.193 | -0.302 | **-0.809** | -0.229 |
| Letter fluency | 0.361 | **0.403** | 0.230 | **0.656** | 0.184 |
| Category fluency | 0.379 | **0.405** | 0.318 | **0.632** | 0.183 |
| SCWT card III time | -0.194 | **-0.465** | -0.362 | **-0.527** | -0.302 |
| Digit span forward span length | 0.273 | 0.090 | -0.010 | 0.169 | **0.902** |
| Digit span forward score | 0.306 | 0.124 | 0.017 | 0.183 | **0.901** |
| Digit span backward score | 0.363 | 0.196 | 0.123 | 0.382 | **0.630** |
| Digit span backward span length | 0.313 | 0.151 | 0.106 | 0.368 | **0.595** |
| RAVLT learning curve | 0.040 | -0.174 | -0.070 | 0.024 | -0.396 |

Variable loads associated with a specific common factor are grouped. Factor loads > 0.4 are in bold.
*Abbreviations:* BNT – Boston Naming Test; BVFDT – Benton Visual Form Discrimination Test; MOCA – Montreal Cognitive Assessment; RAVLT – Rey Auditory Verbal Learning Test; RCFT – Rey Complex Figure Test; SCWT – Stroop Color Word Test; ToL – Tower of London

### 7 Exploratory univariate linear mixed-model analyses of neuropsychological test outcomes

**Table S5:** Group differences from the exploratory univariate linear mixed-model analyses on the individual neuropsychological test outcomes for the crude and adjusted models in the intention-to-treat sample.

| | Baseline | | T1 | | Group difference (crude model) | | | | Group difference (adjusted model)* | | | |
|---|---|---|---|---|---|---|---|---|---|---|---|---|
| | Active control M (SD) | Cognitive training M (SD) | Active control M (SD) | Cognitive training M (SD) | B [SE] | 95% CI | *P* value (FDR) | *P* value (raw) | B [SE] | 95% CI | *P* value (FDR) | *P* value (raw) |
| ToL S1 accuracy (%, N=126) | 96.5 (5.1) | 96.0 (5.9) | 96.9 (4.9) | 96.2 (4.7) | -0.54 [0.81] | -2.13 to 1.06 | 0.824 | 0.505 | -0.57 [0.81] | -2.17 to 1.02 | 0.827 | 0.480 |
| ToL S2 accuracy (%, N=126) | 92.0 (8.7) | 90.6 (9.1) | 92.5 (8.5) | 90.6 (11.6) | -1.66 [1.78] | -5.19 to 1.87 | 0.759 | 0.354 | -1.29 [1.81] | -4.87 to 2.28 | 0.827 | 0.475 |
| ToL S3 accuracy (%, N=126) | 87.7 (10.9) | 86.8 (12.0) | 88.8 (9.1) | 89.0 (13.2) | 0.51 [1.89] | -3.23 to 4.25 | 0.907 | 0.790 | 0.06 [1.92] | -3.75 to 3.87 | 0.974 | 0.974 |
| ToL S4 accuracy (%, N=126) | 75.9 (13.5) | 75.0 (15.2) | 80.8 (14.1) | 78.7 (17.2) | -1.74 [2.58] | -6.83 to 3.36 | 0.824 | 0.501 | -1.61 [2.60] | -6.75 to 3.53 | 0.830 | 0.536 |
| ToL S5 accuracy (%, N=126) | 59.7 (20.4) | 57.6 (20.0) | 65.3 (21.4) | 65.3 (22.2) | 1.54 [2.85] | -4.10 to 7.18 | 0.871 | 0.590 | 0.25 [2.82] | -5.34 to 5.83 | 0.961 | 0.930 |
| ToL S1 reaction time (s, N=126) | 6.4 (1.9) | 5.9 (1.8) | 5.9 (2.1) | 5.3 (1.6) | -0.30 [0.24] | -0.78 to 0.17 | 0.759 | 0.209 | -0.18 [0.24] | -0.66 to 0.29 | 0.827 | 0.445 |
| ToL S2 reaction time (s, N=126) | 8.3 (2.6) | 8.1 (2.9) | 7.4 (2.1) | 7.1 (2.4) | -0.22 [0.25] | -0.71 to 0.28 | 0.759 | 0.392 | -0.15 [0.25] | -0.65 to 0.35 | 0.830 | 0.562 |
| ToL S3 reaction time (s, N=126) | 11.0 (2.7) | 11.0 (3.5) | 10.6 (3.3) | 10.0 (3.5) | -0.61 [0.38] | -1.37 to 0.14 | 0.574 | 0.108 | -0.56 [0.39] | -1.32 to 0.20 | 0.775 | 0.150 |
| ToL S4 reaction time (s, N=126) | 15.6 (3.7) | 15.6 (4.8) | 15.2 (4.8) | 13.8 (4.7) | -1.46 [0.60] | -2.64 to -0.28 | 0.233 | **0.015** | -1.23 [0.60] | -2.41 to -0.05 | 0.496 | **0.042** |
| ToL S5 reaction time (s, N=125) | 22.0 (4.9) | 21.5 (4.8) | 21.5 (4.3) | 20.9 (4.5) | -0.30 [0.68] | -1.64 to 1.05 | 0.907 | 0.663 | -0.07 [0.68] | -1.42 to 1.28 | 0.961 | 0.914 |
| MoCA (N=133) | 26.0 (2.3) | 26.4 (1.9) | 26.0 (2.2) | 26.2 (2.3) | 0.04 [0.35] | -0.66 to 0.74 | 0.918 | 0.918 | -0.10 [0.35] | -0.78 to 0.59 | 0.961 | 0.781 |
| SCWT card I (s, N=133) | 56.3 (15.1) | 54.1 (10.7) | 54.3 (13.2) | 50.8 (9.1) | -2.17 [1.35] | -4.85 to 0.51 | 0.574 | 0.111 | -1.57 [1.35] | -4.25 to 1.07 | 0.823 | 0.239 |
| SCWT card II (s, N=133) | 68.0 (13.8) | 68.2 (18) | 68.6 (16.4) | 64.8 (15.8) | -3.95 [1.83] | -7.57 to -0.34 | 0.331 | **0.032** | -3.64 [1.82] | -7.25 to -0.03 | 0.496 | **0.048** |
| SCWT card III (s, N=132) | 106.9 (33.9) | 114.9 (49.3) | 103.4 (30.7) | 101.7 (28.6) | -6.02 [3.33] | -12.61 to 0.56 | 0.566 | 0.073 | -5.65 [3.38] | -12.33 to 1.03 | 0.601 | 0.097 |
| SCWT interference score (N=132)# | 1.6 (0.3) | 1.7 (0.6) | 1.5 (0.2) | 1.6 (0.2) | 0.03 [0.03] | -0.03 to 0.09 | 0.759 | 0.377 | 0.02 [0.03] | -0.04 to 0.08 | 0.827 | 0.473 |
| Letter fluency (N=133) | 38.8 (11.9) | 38 (10.6) | 42.6 (12.5) | 42.5 (12.7) | 0.65 [1.16] | -1.66 to 2.95 | 0.871 | 0.580 | 0.25 [1.18] | -2.08 to 2.58 | 0.961 | 0.829 |
| Category fluency (N=132) | 23.6 (5.4) | 22.2 (5.1) | 23.6 (6.1) | 21.8 (5.7) | -0.81 [0.79] | -2.38 to 0.76 | 0.759 | 0.311 | -0.98 [0.79] | -2.55 to 0.59 | 0.823 | 0.219 |
| RCFT (N=133) | 30.2 (3.3) | 31.0 (3.6) | 31.0 (3.5) | 30.5 (3.8) | -1.13 [0.46] | -2.04 to -0.22 | 0.233 | **0.015** | -1.32 [0.46] | -2.24 to -0.41 | 0.155 | **0.005** |
| RAVLT learning curve (N=133)† | 0.26 (0.12) | 0.30 (0.10) | 0.30 (0.10) | 0.30 (0.10) | -0.02 [0.02] | -0.06 to 0.03 | 0.759 | 0.416 | -0.02 [0.02] | -0.06 to 0.03 | 0.827 | 0.454 |
| RAVLT direct recall (N=133) | 41.9 (11.4) | 41.2 (10.8) | 46.4 (11.5) | 45.6 (10.4) | -0.24 [1.24] | -2.69 to 2.21 | 0.912 | 0.849 | -0.52 [1.26] | -3.00 to 1.97 | 0.919 | 0.682 |
| RAVLT delayed recall (N=133) | 8.3 (3.5) | 8.3 (3.5) | 9.5 (3.3) | 9.9 (3.1) | 0.34 [0.40] | -0.45 to 1.14 | 0.759 | 0.395 | 0.29 [0.40] | -0.51 to 1.09 | 0.827 | 0.473 |
| RAVLT recognition (N=133) | 28.5 (2.1) | 28.4 (2.0) | 28.5 (1.9) | 28.4 (1.9) | -0.11 [0.29] | -0.69 to 0.47 | 0.907 | 0.719 | -0.19 [0.30] | -0.77 to 0.40 | 0.830 | 0.529 |
| Digit span forward score (N=133) | 9.5 (1.8) | 9.4 (1.6) | 9.5 (1.9) | 9.4 (1.7) | 0.02 [0.24] | -0.45 to 0.50 | 0.918 | 0.917 | -0.07 [0.24] | -0.55 to 0.41 | 0.961 | 0.772 |
| Digit span forward span length (N=133) | 6.2 (1.0) | 6.2 (1.0) | 6.2 (1.0) | 6.2 (1.0) | -0.05 [0.15] | -0.34 to 0.23 | 0.907 | 0.709 | -0.12 [0.14] | -0.41 to 0.16 | 0.827 | 0.393 |
| Digit span backward score (N=133) | 7.3 (1.6) | 6.9 (1.6) | 7.4 (1.8) | 7 (1.6) | -0.22 [0.24] | -0.70 to 0.27 | 0.759 | 0.378 | -0.23 [0.24] | -0.71 to 0.24 | 0.827 | 0.332 |
| Digit span backward span length (N=133) | 5.1 (1.0) | 4.9 (1.1) | 5.3 (1.1) | 5 (0.8) | -0.23 [0.15] | -0.53 to 0.08 | 0.633 | 0.143 | -0.27 [0.15] | -0.57 to 0.04 | 0.601 | 0.083 |
| LLT learning curve (N=132) | 0.61 (0.31) | 0.7 (0.30) | 0.6 (0.30) | 0.6 (0.30) | 0.01 [0.04] | -0.07 to 0.09 | 0.912 | 0.853 | 0.01 [0.04] | -0.08 to 0.09 | 0.961 | 0.874 |
| LLT direct recall (N=132) | 20.9 (16.2) | 21.2 (21.8) | 21.9 (19.3) | 22.8 (16.8) | 0.75 [2.69] | -4.57 to 6.07 | 0.907 | 0.782 | 0.38 [2.64] | -4.85 to 5.60 | 0.961 | 0.887 |
| LLT delayed recall (N=130) | 1.8 (3.0) | 1.8 (4.7) | 1.6 (3.0) | 1.4 (3.0) | -0.13 [0.48] | -1.09 to 0.83 | 0.907 | 0.784 | -0.26 [0.48] | -1.20 to 0.68 | 0.833 | 0.591 |
| BNT (N=133) | 55.5 (3.1) | 55.4 (3.5) | 56.5 (2.6) | 56.2 (3.2) | -0.30 [0.28] | -0.85 to 0.26 | 0.759 | 0.291 | -0.35 [0.29] | -0.92 to 0.22 | 0.823 | 0.228 |
| BVFDT (N=132) | 29.9 (2.5) | 30.1 (1.8) | 29.9 (2.5) | 30.3 (2.5) | 0.37 [0.41] | -0.45 to 1.18 | 0.759 | 0.376 | 0.46 [0.41] | -0.36 to 1.28 | 0.827 | 0.271 |

Significant differences are marked in bold. *Corrected for age, sex and education in years; #Stroop interference score calculation: Stroop card III time / Stroop card II time[30]; †RAVLT learning curve calculated similar to the LLT learning curve model.[9] *Abbreviations:* BNT – Boston Naming Test; BVFDT – Benton Visual Form Discrimination Test; MoCA – Montreal Cognitive Assessment; RAVLT – Rey Auditory Verbal Learning Test; RCFT – Rey Complex Figure Test; SCWT – Stroop Color Word Test; S1-S5 – ToL difficulty load 1-5; ToL – Tower of London.

### 8 Improvement on the experimental and active control condition

**Table S6:** Results from the linear mixed-model analyses of the association between overall training session (1-24) or categorized training sessions, and standardized overall training score or separate training component score in the CT group in the intention-to-treat sample.

| | **Overall session effect** | | | **Phase II vs. phase I** Session 5-8 vs. 1-4 | | | **Phase III vs. phase II** Session 9-12 vs. 5-8 | | | **Phase IV vs. phase III** Session 13-16 vs. 9-12 | | | **Phase V vs. phase IV** Session 17-20 vs. 13-16 | | | **Phase VI vs. phase V** Session 21-24 vs. 17-20 | | |
|---|---|---|---|---|---|---|---|---|---|---|---|---|---|---|---|---|---|---|
| | B [SE] | 95% CI | *P* value | B [SE] | 95% CI | *P* value | B [SE] | 95% CI | *P* value | B [SE] | 95% CI | *P* value | B [SE] | 95% CI | *P* value | B [SE] | 95% CI | *P* value |
| **Cognitive training** | | | | | | | | | | | | | | | | | | |
| Overall training | **0.028 [0.001]** | **0.026 to 0.030** | **< 0.001** | **0.238 [0.030]** | **0.179 to 0.296** | **< 0.001** | **0.140 [0.029]** | **0.082 to 0.197** | **< 0.001** | **0.112 [0.030]** | **0.054 to 0.170** | **0.002** | 0.051 [0.030] | -0.007 to 0.110 | 0.310 | 0.032 [0.030] | -0.027 to 0.091 | 0.642 |
| 1 Drumsolo | **0.016 [0.003]** | **0.011 to 0.022** | **< 0.001** | 0.094 [0.088] | -0.081 to 0.268 | 0.291 | 0.062 [0.088] | -0.112 to 0.237 | 0.481 | 0.156 [0.089] | -0.019 to 0.332 | 0.139 | -0.088 [0.089] | -0.262 to 0.087 | 0.506 | 0.105 [0.089] | -0.069 to 0.280 | 0.642 |
| 2 Shopshift | **0.033 [0.003]** | **0.027 to 0.038** | **< 0.001** | **0.159 [0.063]** | **0.035 to 0.284** | **0.015** | **0.177 [0.062]** | **0.054 to 0.300** | **0.011** | 0.137 [0.063] | 0.013 to 0.261 | 0.072 | 0.091 [0.062] | -0.032 to 0.214 | 0.318 | 0.043 [0.064] | -0.083 to 0.169 | 0.642 |
| 3 Birdz | **0.035 [0.003]** | **0.030 to 0.041** | **< 0.001** | **0.473 [0.069]** | **0.337 to 0.610** | **< 0.001** | **0.197 [0.067]** | **0.065 to 0.330** | **0.010** | 0.157 [0.068] | 0.022 to 0.291 | 0.072 | 0.008 [0.069] | -0.129 to 0.144 | 0.983 | -0.055 [0.070] | -0.193 to 0.083 | 0.642 |
| 4 Totem | **0.031 [0.003]** | **0.026 to 0.037** | **< 0.001** | **0.208 [0.079]** | **0.051 to 0.364** | **0.012** | 0.149 [0.080] | -0.008 to 0.306 | 0.087 | 0.067 [0.080] | -0.090 to 0.224 | 0.469 | 0.127 [0.079] | -0.029 to 0.283 | 0.310 | 0.078 [0.081] | -0.081 to 0.237 | 0.642 |
| 5 Flip | **0.016 [0.003]** | **0.011 to 0.022** | **< 0.001** | **0.216 [0.049]** | **0.120 to 0.311** | **< 0.001** | **0.114 [0.048]** | **0.019 to 0.208** | **0.029** | 0.005 [0.048] | -0.090 to 0.100 | 0.915 | 0.034 [0.049] | -0.063 to 0.131 | 0.687 | -0.060 [0.050] | -0.158 to 0.038 | 0.642 |
| 6 Line-up | **0.016 [0.003]** | **0.011 to 0.022** | **< 0.001** | **0.178 [0.057]** | **0.066 to 0.290** | **0.003** | **0.156 [0.056]** | **0.046 to 0.266** | **0.011** | 0.030 [0.057] | -0.082 to 0.141 | 0.645 | -0.001 [0.057] | -0.113 to 0.110 | 0.983 | -0.018 [0.057] | -0.131 to 0.094 | 0.838 |
| 7 Pengo | **0.045 [0.003]** | **0.039 to 0.050** | **< 0.001** | **0.368 [0.068]** | **0.234 to 0.502** | **< 0.001** | **0.213 [0.064]** | **0.087 to 0.340** | **0.004** | **0.200 [0.065]** | **0.072 to 0.328** | **0.016** | 0.088 [0.068] | -0.046 to 0.221 | 0.345 | 0.061 [0.067] | -0.072 to 0.193 | 0.642 |
| 8 Bunny | **0.015 [0.003]** | **0.010 to 0.021** | **< 0.001** | **0.206 [0.053]** | **0.101 to 0.310** | **< 0.001** | 0.059 [0.052] | -0.044 to 0.162 | 0.303 | 0.071 [0.053] | -0.034 to 0.176 | 0.259 | -0.001 [0.054] | -0.107 to 0.104 | 0.983 | 0.001 [0.054] | -0.106 to 0.108 | 0.992 |
| 9 N-back | **0.037 [0.003]** | **0.032 to 0.043** | **< 0.001** | **0.212 [0.069]** | **0.077 to 0.347** | **0.003** | **0.184 [0.068]** | **0.050 to 0.318** | **0.013** | 0.149 [0.069] | 0.014 to 0.284 | 0.072 | 0.119 [0.070] | -0.018 to 0.255 | 0.310 | 0.020 [0.070] | -0.118 to 0.158 | 0.838 |
| 10 Mentile | **0.025 [0.003]** | **0.019 to 0.030** | **< 0.001** | **0.194 [0.057]** | **0.082 to 0.305** | **0.001** | 0.098 [0.055] | -0.011 to 0.207 | 0.098 | 0.095 [0.056] | -0.015 to 0.205 | 0.139 | 0.079 [0.056] | -0.031 to 0.189 | 0.318 | 0.078 [0.057] | -0.033 to 0.189 | 0.642 |
| 11 Square logic | **0.046 [0.003]** | **0.041 to 0.052** | **< 0.001** | **0.194 [0.065]** | **0.065 to 0.322** | **0.005** | **0.250 [0.063]** | **0.125 to 0.374** | **0.001** | **0.171 [0.064]** | **0.045 to 0.298** | **0.039** | **0.233 [0.065]** | **0.105 to 0.361** | **0.006** | 0.056 [0.065] | -0.072 to 0.184 | 0.642 |
| 12 Bait | **0.034 [0.003]** | **0.029 to 0.040** | **< 0.001** | **0.298 [0.053]** | **0.194 to 0.403** | **< 0.001** | **0.192 [0.051]** | **0.092 to 0.292** | **0.001** | 0.102 [0.051] | 0.002 to 0.202 | 0.093 | 0.092 [0.051] | -0.008 to 0.193 | 0.310 | 0.077 [0.052] | -0.025 to 0.179 | 0.642 |
| 13 Fuzzle | **0.011 [0.003]** | **0.005 to 0.017** | **< 0.001** | **0.238 [0.115]** | **0.009 to 0.466** | **0.045** | -0.085 [0.108] | -0.298 to 0.128 | 0.463 | 0.131 [0.106] | -0.079 to 0.342 | 0.279 | -0.018 [0.107] | -0.230 to 0.195 | 0.983 | -0.073 [0.100] | -0.271 to 0.126 | 0.642 |
| **Active control** | | | | | | | | | | | | | | | | | | |
| Overall active control | **0.022 [0.002]** | **0.019 to 0.025** | **< 0.001** | 0.213 [0.051] | 0.112 to 0.313 | 0.138 | -0.003 [0.052] | -0.105 to 0.099 | 0.759 | 0.132 [0.052] | 0.029 to 0.235 | 0.092 | 0.063 [0.052] | -0.038 to 0.165 | 0.661 | 0.052 [0.055] | -0.056 to 0.161 | 0.920 |
| 1 Solitaire | **0.012 [0.003]** | **0.007 to 0.017** | **< 0.001** | **0.114 [0.077]** | **-0.037 to 0.265** | **< 0.001** | -0.044 [0.078] | -0.197 to 0.109 | 0.474 | 0.142 [0.078] | -0.011 to 0.294 | 0.628 | 0.042 [0.076] | -0.108 to 0.192 | 0.661 | -0.008 [0.082] | -0.170 to 0.154 | 0.550 |
| 2 Hangman | **0.012 [0.003]** | **0.007 to 0.017** | **< 0.001** | **0.195 [0.024]** | **0.148 to 0.242** | **< 0.001** | 0.038 [0.024] | -0.010 to 0.086 | 0.699 | **0.012 [0.025]** | **-0.036 to 0.060** | **< 0.001** | **0.011 [0.024]** | **-0.037 to 0.059** | **0.026** | **0.021 [0.026]** | **-0.030 to 0.072** | **0.043** |
| 3 Trivia | **0.043 [0.003]** | **0.038 to 0.048** | **< 0.001** | **0.352 [0.051]** | **0.252 to 0.451** | **< 0.001** | -0.049 [0.052] | -0.151 to 0.054 | 0.951 | **0.273 [0.053]** | **0.170 to 0.377** | **0.024** | 0.142 [0.052] | 0.040 to 0.243 | 0.444 | 0.140 [0.054] | 0.032 to 0.247 | 0.550 |

*P* values are FDR-corrected values. Significant differences are marked in bold.

### 9 Association between CT and active control improvement and executive function change

**Table S7a:** Association between the improvement on the overall CT and active control, and specific CT and control games, and the Tower of London outcomes.

| Training component | Tower of London - accuracy S1-S5 | | | Tower of London - reaction time S1-S5 | | | Tower of London - reaction time S4 | | |
|---|---|---|---|---|---|---|---|---|---|
| | B [SE] | 95% CI | *P* value | B [SE] | 95% CI | *P* value | B [SE] | 95% CI | *P* value |
| **Cognitive training** | | | | | | | | | |
| Overall training | **10.788 [4.659]** | **1.464 to 20.112** | **0.024** | -6.308 [3.248] | -12.816 to 0.200 | 0.057 | -4.718 [3.840] | -12.331 to 2.895 | 0.222 |
| 1 Drumsolo | 4.126 [2.338] | -0.554 to 8.806 | 0.083 | -0.303 [1.633] | -3.578 to 2.972 | 0.854 | 0.985 [1.914] | -2.812 to 4.782 | 0.608 |
| 2 Shopshift | **4.875 [2.167]** | **0.538 to 9.213** | **0.028** | **-3.830 [1.504]** | **-6.840 to -0.820** | **0.014** | -3.359 [1.773] | -6.871 to 0.154 | 0.061 |
| 3 Birdz | -1.543 [3.095] | -7.738 to 4.653 | 0.620 | 0.267 [2.107] | -3.958 to 4.492 | 0.900 | 3.120 [2.464] | -1.769 to 8.008 | 0.208 |
| 4 Totem | **4.948 [1.935]** | **1.075 to 8.821** | **0.013** | **-2.997 [1.332]** | **-5.667 to -0.327** | **0.029** | -2.684 [1.574] | -5.805 to 0.438 | 0.091 |
| 5 Flip | 1.523 [3.859] | -6.203 to 9.248 | 0.695 | -0.907 [2.621] | -6.164 to 4.350 | 0.731 | 0.559 [3.073] | -5.538 to 6.657 | 0.856 |
| 6 Line-up | 3.548 [4.316] | -5.092 to 12.188 | 0.414 | -4.669 [2.891] | -10.466 to 1.129 | 0.112 | -3.669 [3.425] | -10.463 to 3.124 | 0.287 |
| 7 Pengo | 3.165 [2.853] | -2.546 to 8.876 | 0.272 | -2.882 [1.953] | -6.794 to 1.030 | 0.146 | -2.965 [2.298] | -7.521 to 1.591 | 0.200 |
| 8 Bunny | 1.260 [3.867] | -6.481 to 9.002 | 0.746 | 0.857 [2.624] | -4.406 to 6.120 | 0.745 | 2.635 [3.058] | -3.431 to 8.701 | 0.391 |
| 9 N-back | **4.673 [2.062]** | **0.546 to 8.799** | **0.027** | -1.858 [1.453] | -4.769 to 1.054 | 0.206 | -2.854 [1.698] | -6.221 to 0.513 | 0.096 |
| 10 Mentile | 2.710 [3.189] | -3.673 to 9.092 | 0.399 | **-4.562 [2.118]** | **-8.808 to -0.317** | **0.036** | -4.909 [2.502] | -9.869 to 0.051 | 0.052 |
| 11 Square logic | **6.189 [2.537]** | **1.111 to 11.267** | **0.018** | -3.284 [1.806] | -6.900 to 0.332 | 0.074 | -2.684 [2.118] | -6.881 to 1.513 | 0.208 |
| 12 Bait | 5.532 [3.560] | -1.593 to 12.656 | 0.126 | -2.479 [2.434] | -7.361 to 2.403 | 0.313 | -2.871 [2.865] | -8.555 to 2.812 | 0.319 |
| 13 Fuzzle | -0.892 [3.052] | -7.002 to 5.218 | 0.771 | 1.105 [2.070] | -3.047 to 5.256 | 0.596 | 2.602 [2.436] | -2.232 to 7.436 | 0.288 |
| **Active control** | | | | | | | | | |
| Overall active control | 6.280 [4.479] | -2.672 to 15.231 | 0.166 | 1.881 [6.067] | -10.253 to 14.016 | 0.758 | 3.559 [6.603] | -9.579 to 16.698 | 0.591 |
| 1 Solitaire | 2.877 [2.144] | -1.409 to 7.162 | 0.185 | 2.055 [2.888] | -3.722 to 7.832 | 0.480 | 2.343 [3.159] | -3.942 to 8.627 | 0.460 |
| 2 Hangman | -1.908 [4.710] | -11.321 to 7.505 | 0.687 | **14.676 [6.132]** | **2.425 to 26.926** | **0.020** | **17.841 [6.672]** | **4.583 to 31.099** | **0.009** |
| 3 Trivia | 2.387 [2.461] | -2.533 to 7.306 | 0.336 | -5.011 [3.267] | -11.544 to 1.522 | 0.130 | -4.802 [3.560] | -11.882 to 2.279 | 0.181 |

Significant associations are marked in bold.

**Table S7b:** Association between the improvement on the overall CT and active control, and specific CT and control games, and the Stroop Color Word Test card II (color-only) and card III (color-word) outcomes.

| Training component | Stroop color word test - card II | | | Stroop color word test - card III | | |
|---|---|---|---|---|---|---|
| | B [SE] | 95% CI | *P* value | B [SE] | 95% CI | *P* value |
| **Cognitive training** | | | | | | |
| Overall training | **-231.651 [72.597]** | **-376.597 to -86.706** | **0.002** | **-507.164 [119.218]** | **-745.191 to -269.137** | **< .001** |
| 1 Drumsolo | -49.892 [36.600] | -122.967 to 23.183 | 0.177 | -91.406 [65.524] | -222.228 to 39.416 | 0.168 |
| 2 Shopshift | **-78.483 [33.714]** | **-145.795 to -11.171** | **0.023** | **-198.288 [58.651]** | **-315.388 to -81.189** | **0.001** |
| 3 Birdz | -68.213 [43.836] | -155.735 to 19.309 | 0.124 | **-193.478 [72.988]** | **-339.204 to -47.752** | **0.010** |
| 4 Totem | -63.256 [30.494] | -124.139 to -2.374 | 0.042 | **-142.671 [53.779]** | **-250.043 to -35.298** | **0.010** |
| 5 Flip | -53.132 [57.714] | -168.362 to 62.099 | 0.361 | -109.234 [103.723] | -316.323 to 97.856 | 0.296 |
| 6 Line-up | **-202.073 [65.377]** | **-332.602 to -71.543** | **0.003** | **-375.896 [107.972]** | **-591.470 to -160.322** | **0.001** |
| 7 Pengo | **-100.421 [41.751]** | **-183.779 to -17.063** | **0.019** | **-208.294 [74.121]** | **-356.282 to -60.306** | **0.007** |
| 8 Bunny | -41.761 [58.025] | -157.612 to 74.090 | 0.474 | -155.818 [102.305] | -360.077 to 48.441 | 0.133 |
| 9 N-back | -59.930 [31.722] | -123.264 to 3.405 | 0.063 | **-116.499 [57.804]** | **-231.907 to -1.090** | **0.048** |
| 10 Mentile | **-150.344 [45.100]** | **-240.390 to -60.299** | **0.001** | **-277.333 [81.654]** | **-440.361 to -114.305** | **0.001** |
| 11 Square logic | -70.104 [39.365] | -148.698 to 8.490 | 0.080 | **-157.588 [69.446]** | **-296.241 to -18.935** | **0.027** |
| 12 Bait | -49.494 [54.847] | -159.000 to 60.012 | 0.370 | **-194.435 [95.228]** | **-384.565 to -4.305** | **0.045** |
| 13 Fuzzle | 8.387 [42.618] | -76.703 to 93.477 | 0.845 | -64.952 [71.235] | -207.177 to 77.273 | 0.365 |
| **Active control** | | | | | | |
| Overall active control | -42.265 [101.838] | -245.480 to 160.950 | 0.679 | -2.808 [153.232] | -308.659 to 303.044 | 0.985 |
| 1 Solitaire | -50.143 [48.421] | -146.767 to 46.480 | 0.304 | 56.733 [74.219] | -91.409 to 204.875 | 0.447 |
| 2 Hangman | 128.017 [106.074] | -83.649 to 339.684 | 0.232 | 105.047 [162.787] | -219.878 to 429.971 | 0.521 |
| 3 Trivia | -7.232 [56.317] | -119.610 to 105.146 | 0.898 | -100.125 [82.167] | -264.132 to 63.882 | 0.227 |

Significant associations are marked in bold.

#### 10 Analysis of the effect of CT on exploratory psychiatric outcomes

Multivariate analysis of the overall psychiatric symptoms (N=130) are shown in Table S9. The CT group experienced on average 0.17 standard deviation less psychiatric symptoms averaged across questionnaires after training compared with the AC group. This difference was not significant: B[SE]: -0.17 [0.09], 95% CI: -0.34 to 0.00, p=0.051 (raw model); B[SE]: -0.15 [0.09], 95% CI: -0.32 to 0.03, p=0.093 (adjusted model). The CT group experienced statistically significant lower impulse control disorder symptoms after training compared with the AC group, which was not significant after correcting for age, sex and years of education. This effect seemed driven by a post-training increase in impulsive behavior in the AC group. Post-hoc QUIP-RS sub-score analysis (see Table S10) showed that this effect was mainly driven by a larger increase on the hyper-sexuality and compulsive buying sub-scale in the AC group and a larger decrease in the hobbyism/punding sub-score in the CT group. Mean scores remained under the respective cut-off scores for clinically relevant symptoms. At follow-up, no group differences were present anymore (see Table S11 and Table S12).

**Table S8:** Group differences on the exploratory psychiatric outcome measures for the crude and adjusted analysis models in the intention-to-treat and per-protocol sample.

| | Baseline | | T1 | | Group difference (crude model) | | | Group difference (adjusted model)* | | |
|---|---|---|---|---|---|---|---|---|---|---|
| | Active control M (SD) | Cognitive training M (SD) | Active control M (SD) | Cognitive training M (SD) | B [SE] | 95% CI | *P* value | B [SE] | 95% CI | *P* value |
| **INTENTION-TO-TREAT SAMPLE** | | | | | | | | | | |
| Overall psychiatric symptoms | | | | | -0.17 [0.09] | -0.34 to 0.00 | 0.051 | -0.15 [0.09] | -0.32 to 0.03 | 0.093 |
| BDI | 7.9 (4.1) | 8.2 (4.0) | 9.2 (4.6) | 8.4 (4.3) | -0.21 [0.13] | -0.47 to 0.05 | 0.116 | -0.18 [0.13] | -0.44 to 0.07 | 0.163 |
| QUIP-RS | 19.2 (12.7) | 15.8 (12.8) | 21.4 (12.1) | 16.0 (10.6) | -0.29 [0.14] | -0.56 to -0.02 | 0.033 | -0.27 [0.14] | -0.54 to -0.00 | 0.052 |
| PAS | 10.5 (6.8) | 10.3 (6.6) | 11.5 (6.6) | 11.2 (7.7) | -0.06 [0.13] | -0.32 to 0.20 | 0.668 | -0.03 [0.13] | -0.29 to 0.23 | 0.800 |
| AS | 13.4 (4.5) | 13.2 (4.5) | 14.0 (5.3) | 13.1 (4.3) | -0.14 [0.13] | -0.40 to 0.12 | 0.295 | -0.12 [0.13] | -0.38 to 0.15 | 0.383 |
| **PER-PROTOCOL SAMPLE** | | | | | | | | | | |
| Overall psychiatric symptoms (N=127) | | | | | -0.12 [0.09] | -0.29 to 0.05 | 0.174 | -0.10 [0.09] | -0.27 to 0.08 | 0.273 |
| BDI | 7.9 (3.8) | 8.1 (3.9) | 8.7 (4.3) | 8.4 (4.4) | -0.11 [0.13] | -0.37 to 0.15 | 0.399 | -0.09 [0.13] | -0.35 to 0.17 | 0.497 |
| QUIP-RS | 18.8 (12.9) | 16.1 (12.9) | 20.8 (12.0) | 16.0 (10.7) | -0.26 [0.14] | -0.53 to 0.00 | 0.053 | -0.24 [0.14] | -0.51 to 0.03 | 0.079 |
| PAS | 10.5 (6.8) | 10.3 (6.8) | 11.1 (6.4) | 11.3 (7.7) | 0.00 [0.13] | -0.26 to 0.26 | 1.000 | -0.02 [0.13] | -0.24 to 0.28 | 0.870 |
| AS | 13.6 (4.4) | 13.4 (4.6) | 14.0 (5.3) | 13.3 (4.3) | -0.10 [0.13] | -0.36 to 0.16 | 0.435 | -0.08 [0.13] | -0.34 to 0.18 | 0.538 |

Negative estimates indicate effects in favor of CT; *Corrected for age, sex and education in years.
Abbreviations: BDI = Beck Depression Inventory; QUIP-RS = Questionnaire for Impulsive-Compulsive Disorders in Parkinson's Disease – Rating Scale; PAS = Parkinson Anxiety Scale; AS = Apathy Scale.

**Table S9:** Post-hoc analysis of group differences on QUIP-RS sub-scales after intervention from the exploratory multivariate linear mixed-model analysis for the crude and adjusted analysis models in the intention-to-treat sample.

| | Baseline | | T1 | | Group difference (crude model) | | | Group difference (adjusted model)* | | |
|---|---|---|---|---|---|---|---|---|---|---|
| | Active control M (SD) | Cognitive training M (SD) | Active control M (SD) | Cognitive training M (SD) | B [SE] | 95% CI | *P* value | B [SE] | 95% CI | *P* value |
| QUIP-RS sub-scale#: | | | | | | | | | | |
| Gambling | 0.29 (0.85) | 0.06 (0.39) | 0.30 (1.11) | 0.08 (0.32) | -0.09 [0.35] | -0.77 to 0.59 | 0.798 | 0.11 [0.34] | -0.57 to 0.78 | 0.755 |
| Sex | 2.85 (2.40) | 2.55 (2.57) | 3.79 (2.97) | 2.97 (2.93) | **-0.77 [0.35]** | **-1.45 to -0.08** | **0.028** | -0.57 [0.34] | -1.24 to 0.10 | 0.096 |
| Buying | 2.03 (2.47) | 1.82 (2.16) | 2.91 (2.74) | 1.80 (2.16) | **-0.99 [0.35]** | **-1.67 to -0.31** | **0.005** | **-0.79 [0.34]** | **-1.46 to -0.12** | **0.021** |
| Eating | 3.57 (2.88) | 3.02 (3.04) | 3.97 (2.99) | 3.26 (2.77) | -0.32 [0.36] | -1.02 to 0.38 | 0.374 | -0.11 [0.35] | -0.81 to 0.58 | 0.754 |
| Hobbyism and punding | 3.42 (2.47) | 2.77 (2.40) | 3.65 (2.40) | 2.39 (1.90) | **-0.87 [0.35]** | **-1.56 to -0.19** | **0.012** | **-0.68 [0.34]** | **-1.35 to -0.01** | **0.047** |
| Compulsive medication use | 3.60 (3.25) | 2.73 (2.83) | 3.33 (3.13) | 3.24 (2.46) | 0.35 [0.35] | -0.34 to 1.03 | 0.319 | 0.54 [0.34] | -0.14 to 1.21 | 0.118 |

Negative estimates indicate effects in favor of CT; Significant differences are marked in bold. *Corrected for age, sex and education in years; #According to Weintraub and colleagues[17]
*Abbreviations*: QUIP-RS = Questionnaire for Impulsive-Compulsive Disorders in Parkinson's Disease – Rating Scale

**Table S10:** Group differences at six-months follow-up (T2) on the exploratory psychiatric outcome measures for the crude and adjusted analysis models in the intention-to-treat and per-protocol sample.

| | T2 | | Group difference (crude model) | | | Group difference (adjusted model)* | | |
|---|---|---|---|---|---|---|---|---|
| | Active control M (SD) | Cognitive training M (SD) | B [SE] | 95% CI | *P* value | B [SE] | 95% CI | *P* value |
| **INTENTION-TO-TREAT SAMPLE** | | | | | | | | |
| Overall psychiatric symptoms | | | -0.04 [0.09] | -0.23 to 0.15 | 0.685 | -0.02 [0.10] | -0.21 to 0.17 | 0.834 |
| BDI | 10.0 (4.9) | 8.9 (4.6) | -0.23 [0.14] | -0.51 to 0.04 | 0.099 | -0.21 [0.14] | -0.49 to 0.06 | 0.129 |
| QUIP-RS | 21.2 (14.0) | 19.8 (13.0) | 0.02 [0.15] | -0.27 to 0.31 | 0.891 | 0.04 [0.15] | -0.25 to 0.33 | 0.790 |
| PAS | 10.8 (7.6) | 11.8 (7.5) | 0.12 [0.14] | -0.16 to 0.40 | 0.406 | 0.14 [0.14] | -0.14 to 0.42 | 0.337 |
| AS | 14.5 (5.5) | 14.0 (4.4) | -0.06 [0.14] | -0.33 to 0.22 | 0.697 | -0.04 [0.14] | -0.32 to 0.24 | 0.796 |
| **PER-PROTOCOL SAMPLE** | | | | | | | | |
| Overall psychiatric symptoms (N=122) | | | 0.01 [0.10] | -0.18 to 0.20 | 0.936 | 0.03 [0.10] | -0.17 to 0.22 | 0.787 |
| BDI | 9.7 (4.8) | 9.0 (4.7) | -0.16 [0.14] | -0.44 to 0.12 | 0.272 | -0.14 [0.14] | -0.42 to 0.14 | 0.334 |
| QUIP-RS | 20.8 (13.8) | 19.5 (13.0) | 0.05 [0.15] | -0.24 to 0.34 | 0.759 | 0.06 [0.15] | -0.23 to 0.36 | 0.668 |
| PAS | 10.3 (7.2) | 11.8 (7.6) | 0.17 [0.14] | -0.11 to 0.46 | 0.225 | 0.19 [0.14] | -0.09 to 0.48 | 0.182 |
| AS | 14.4 (5.6) | 14.0 (4.4) | -0.03 [0.14] | -0.31 to 0.25 | 0.840 | -0.01 [0.14] | -0.29 to 0.27 | 0.942 |

Negative estimates indicate effects in favor of CT; *Corrected for age, sex and education in years.
Abbreviations: BDI = Beck Depression Inventory; QUIP-RS = Questionnaire for Impulsive-Compulsive Disorders in Parkinson's Disease – Rating Scale; PAS = Parkinson Anxiety Scale; AS = Apathy Scale.

**Table S11:** Post-hoc analysis of group differences at six-months follow-up (T2) on QUIP-RS sub-scales after intervention from the exploratory multivariate linear mixed-model analysis for the crude and adjusted analysis models in the intention-to-treat sample.

| | T2 | | Group difference (crude model) | | | Group difference (adjusted model)* | | |
|---|---|---|---|---|---|---|---|---|
| | Active control M (SD) | Cognitive training M (SD) | B [SE] | 95% CI | *P* value | B [SE] | 95% CI | *P* value |
| QUIP-RS sub-scale[#]: | | | | | | | | |
| Gambling | 0.33 (1.22) | 0.20 (0.81) | 0.06 [0.34] | -0.61 to 0.74 | 0.852 | 0.27 [0.34] | -0.39 to 0.94 | 0.417 |
| Sex | 3.56 (2.87) | 3.62 (3.46) | 0.10 [0.34] | -0.57 to 0.78 | 0.765 | 0.31 [0.34] | -0.35 to 0.97 | 0.354 |
| Buying | 2.58 (2.76) | 2.54 (2.31) | -0.02 [0.34] | -0.70 to 0.66 | 0.953 | 0.19 [0.34] | -0.47 to 0.85 | 0.573 |
| Eating | 3.70 (2.94) | 3.58 (3.13) | 0.06 [0.35] | -0.63 to 0.75 | 0.863 | 0.28 [0.35] | -0.40 to 0.96 | 0.420 |
| Hobbyism and punding | 3.48 (2.77) | 3.32 (2.43) | 0.07 [0.34] | -0.60 to 0.75 | 0.831 | 0.28 [0.34] | -0.38 to 0.94 | 0.405 |
| Compulsive medication use | 3.77 (3.14) | 3.37 (2.85) | 0.29 [0.34] | -0.39 to 0.97 | 0.399 | 0.50 [0.34] | -0.17 to 1.16 | 0.141 |

Negative estimates indicate effects in favor of CT; *Corrected for age, sex and education in years; [#]According to Weintraub and colleagues[17]
Abbreviations: QUIP-RS = Questionnaire for Impulsive-Compulsive Disorders in Parkinson's Disease – Rating Scale

### 11 Analyses of the neuropsychological and clinical measures at six-months follow-up in the intention-to-treat sample

**Table S12:** Group differences at six-months follow-up (T2) from the multivariate linear mixed-model analyses on the primary and secondary outcome measures for the crude and adjusted analysis models in the intention-to-treat sample.

| | | T2 | | Group difference (crude model) | | | Group difference (adjusted model)* | | |
|---|---|---|---|---|---|---|---|---|---|
| | | Active control M (SD) | Cognitive training M (SD) | B [SE] | 95% CI | *P* value | B [SE] | 95% CI | *P* value |
| Overall ToL accuracy (%) | | 84.5 (9.1) | 84.3 (10.4) | 0.039 [0.106] | -0.169 to 0.248 | 0.711 | 0.027 [0.103] | -0.178 to 0.231 | 0.798 |
| Sub-score (%): | S1 | 96.1 (4.7) | 97.1 (4.7) | 0.191 [0.164] | -0.131 to 0.514 | 0.244 | 0.179 [0.163] | -0.140 to 0.498 | 0.272 |
| | S2 | 92.5 (8.5) | 93.5 (7.3) | 0.121 [0.164] | -0.201 to 0.444 | 0.460 | 0.107 [0.163] | -0.212 to 0.427 | 0.510 |
| | S3 | 88.0 (11.2) | 89.0 (11.7) | 0.053 [0.164] | -0.270 to 0.375 | 0.749 | 0.040 [0.163] | -0.279 to 0.360 | 0.804 |
| | S4 | 77.4 (16.0) | 78.2 (16.5) | 0.015 [0.164] | -0.307 to 0.338 | 0.927 | 0.003 [0.163] | -0.316 to 0.322 | 0.986 |
| | S5 | 68.3 (19.6) | 64.6 (21.4) | -0.185 [0.164] | -0.507 to 0.138 | 0.261 | -0.198 [0.163] | -0.517 to 0.122 | 0.225 |
| **Secondary outcome measures** | | | | | | | | | |
| Overall ToL reaction time (s)# | | 11.5 (3.0) | 11.0 (2.7) | 0.000 [0.097] | -0.193 to 0.192 | 0.998 | 0.054 [0.095] | -0.134 to 0.242 | 0.574 |
| Sub-score (s): | S1 | 5.8 (2.0) | 5.3 (1.8) | -0.069 [0.126] | -0.316 to 0.179 | 0.587 | -0.017 [0.124] | -0.261 to 0.227 | 0.892 |
| | S2 | 7.5 (2.5) | 6.9 (2.5) | -0.085 [0.126] | -0.333 to 0.162 | 0.498 | -0.031 [0.124] | -0.275 to 0.212 | 0.800 |
| | S3 | 10.1 (3.1) | 9.4 (2.8) | -0.083 [0.126] | -0.330 to 0.165 | 0.512 | -0.028 [0.124] | -0.271 to 0.216 | 0.823 |
| | S4 | 14.0 (4.3) | 13.8 (4.5) | 0.054 [0.126] | -0.193 to 0.302 | 0.666 | 0.109 [0.124] | -0.134 to 0.353 | 0.378 |
| | S5 | 20.2 (5.0) | 20.3 (4.8) | 0.183 [0.126] | -0.065 to 0.431 | 0.148 | 0.237 [0.124] | -0.008 to 0.481 | 0.058 |
| Overall subjective cognitive complaints | | | | -0.04 [0.12] | -0.27 to 0.19 | 0.748 | 0.00 [0.12] † | -0.24 to 0.23 | 0.972 |
| Domain: | PD-CFRS | 8.4 (4.9) | 7.2 (4.1) | -0.08 [0.15] | -0.37 to 0.21 | 0.580 | -0.05 [0.15] † | -0.34 to 0.24 | 0.744 |
| | PD-CFRS inf. | 5.4 (4.2) | 5.4 (4.1) | -0.02 [0.18] | -0.37 to 0.33 | 0.902 | 0.02 [0.18] † | -0.33 to 0.37 | 0.912 |
| | CFQ | 37.7 (12.4) | 37.3 (12.4) | 0.00 [0.15] | -0.28 to 0.29 | 0.984 | 0.03 [0.15] † | -0.26 to 0.32 | 0.834 |
| Overall cognitive factors | | | | 0.026 [0.065] | -0.102 to 0.154 | 0.686 | 0.000 [0.066] | -0.128 to 0.129 | 0.995 |
| Factors: | Factor 1 | 0.201 (0.966) | 0.172 (0.890) | -0.124 [0.145] | -0.409 to 0.160 | 0.390 | -0.149 [0.144] | -0.432 to 0.134 | 0.301 |
| | Factor 2 | -0.142 (1.092) | 0.126 (0.829) | 0.170 [0.145] | -0.114 to 0.454 | 0.240 | 0.146 [0.144] | -0.137 to 0.429 | 0.313 |
| | Factor 3 | 0.096 (0.753) | 0.044 (0.795) | -0.039 [0.145] | -0.323 to 0.245 | 0.788 | -0.066 [0.144] | -0.349 to 0.217 | 0.648 |
| | Factor 4 | -0.047 (0.925) | 0.082 (0.957) | 0.125 [0.145] | -0.159 to 0.409 | 0.387 | 0.099 [0.144] | -0.184 to 0.382 | 0.493 |
| | Factor 5 | -0.044 (1.090) | -0.083 (1.113) | 0.000 [0.145] | -0.284 to 0.284 | 1.000 | -0.027 [0.144] | -0.310 to 0.256 | 0.850 |

*Corrected for age, sex and education in years; #Reaction time of correct responses; †Model additionally corrected for credibility/expectancy questionnaire score.
Abbreviations: CFQ = Cognitive Failures Questionnaire; inf. = informant version; PD-CFRS = Parkinson's disease – Cognitive Functional Rating Scale; ToL = Tower of London.

### 12 Analyses of the neuropsychological and clinical measures at six-months follow-up in the per-protocol sample

**Table S13:** Group differences at six-months follow-up (T2) from the multivariate linear mixed-model analyses on the primary and secondary outcome measures for the crude and adjusted analysis models in the per-protocol sample.

| | | **T2** | | **Group difference (crude model)** | | | **Group difference (adjusted model)*** | | |
|---|---|---|---|---|---|---|---|---|---|
| | | Active control M (SD) | Cognitive training M (SD) | B [SE] | 95% CI | *P* value | B [SE] | 95% CI | *P* value |
| **Primary outcome measure** | | | | | | | | | |
| Overall ToL accuracy (%, N=115) | | 84.6 (9.2) | 84.0 (10.3) | 0.048 [0.106] | -0.162 to 0.258 | 0.652 | 0.033 [0.104] | -0.173 to 0.238 | 0.754 |
| Sub-score (%): | S1 | 96.2 (4.6) | 97.1 (4.7) | 0.171 [0.164] | -0.151 to 0.493 | 0.299 | 0.156 [0.163] | -0.163 to 0.476 | 0.336 |
| | S2 | 92.7 (8.5) | 93.4 (7.3) | 0.113 [0.164] | -0.209 to 0.435 | 0.491 | 0.097 [0.163] | -0.222 to 0.416 | 0.551 |
| | S3 | 87.9 (11.3) | 88.9 (11.7) | 0.089 [0.164] | -0.233 to 0.411 | 0.586 | 0.075 [0.163] | -0.245 to 0.394 | 0.647 |
| | S4 | 77.5 (16.4) | 78.0 (16.6) | 0.042 [0.164] | -0.280 to 0.364 | 0.800 | 0.027 [0.163] | -0.293 to 0.346 | 0.870 |
| | S5 | 68.6 (19.3) | 64.1 (21.2) | -0.176 [0.164] | -0.498 to 0.146 | 0.284 | -0.191 [0.163] | -0.511 to 0.128 | 0.240 |
| **Secondary outcome measures** | | | | | | | | | |
| Overall ToL reaction time (s, N=115)# | | 11.5 (2.9) | 11.1 (2.7) | -0.007 [0.097] | -0.199 to 0.184 | 0.941 | 0.041 [0.095] | -0.148 to 0.229 | 0.669 |
| Sub-score (s): | S1 | 5.8 (2.0) | 5.3 (1.8) | -0.076 [0.126] | -0.324 to 0.172 | 0.547 | -0.030 [0.125] | -0.275 to 0.215 | 0.810 |
| | S2 | 7.4 (2.5) | 6.9 (2.5) | -0.080 [0.126] | -0.328 to 0.168 | 0.525 | -0.032 [0.125] | -0.277 to 0.213 | 0.797 |
| | S3 | 10.0 (3.1) | 9.5 (2.8) | -0.094 [0.126] | -0.341 to 0.154 | 0.458 | -0.044 [0.125] | -0.289 to 0.201 | 0.723 |
| | S4 | 13.9 (4.0) | 13.9 (4.4) | 0.053 [0.126] | -0.195 to 0.300 | 0.676 | 0.102 [0.125] | -0.143 to 0.347 | 0.414 |
| | S5 | 20.3 (4.9) | 20.4 (4.8) | 0.162 [0.126] | -0.086 to 0.410 | 0.200 | 0.210 [0.125] | -0.036 to 0.456 | 0.093 |
| Overall subjective cognitive complaints (N=122) | | | | -0.03 [0.12] | -0.26 to 0.20 | 0.780 | -0.02 [0.12]† | -0.25 to 0.22 | 0.889 |
| Domain: | PD-CFRS | 8.4 (4.8) | 7.2 (4.1) | -0.09 [0.15] | -0.37 to 0.20 | 0.540 | -0.07 [0.15]† | -0.37 to 0.22 | 0.616 |
| | PD-CFRS inf. | 5.4 (4.3) | 5.4 (4.2) | -0.01 [0.18] | -0.36 to 0.34 | 0.969 | 0.02 [0.18]† | -0.33 to 0.37 | 0.913 |
| | CFQ | 37.6 (12.1) | 37.4 (12.3) | 0.02 [0.15] | -0.27 to 0.30 | 0.914 | 0.03 [0.15]† | -0.27 to 0.32 | 0.865 |
| Overall cognitive factors (N = 115) | | | | 0.019 [0.066] | -0.109 to 0.148 | 0.766 | -0.003 [0.066] | -0.132 to 0.127 | 0.966 |
| Factors: | Factor 1 | 0.207 (0.982) | 0.167 (0.898) | -0.116 [0.145] | -0.401 to 0.169 | 0.425 | -0.137 [0.145] | -0.421 to 0.147 | 0.343 |
| | Factor 2 | -0.088 (1.052) | 0.111 (0.829) | 0.139 [0.145] | -0.146 to 0.424 | 0.339 | 0.117 [0.145] | -0.167 to 0.402 | 0.417 |
| | Factor 3 | 0.076 (0.758) | 0.040 (0.801) | -0.033 [0.145] | -0.318 to 0.252 | 0.820 | -0.056 [0.145] | -0.340 to 0.228 | 0.700 |
| | Factor 4 | -0.055 (0.940) | 0.070 (0.961) | 0.105 [0.145] | -0.180 to 0.390 | 0.471 | 0.082 [0.145] | -0.202 to 0.367 | 0.569 |
| | Factor 5 | -0.046 (1.086) | -0.094 (1.120) | 0.003 [0.145] | -0.283 to 0.288 | 0.985 | -0.021 [0.145] | -0.305 to 0.263 | 0.885 |

*Corrected for age, sex and education in years; #Reaction time of correct responses; †Model additionally corrected for credibility/expectancy questionnaire score.
Abbreviations: CFQ = Cognitive Failures Questionnaire; inf. = informant version; PD-CFRS = Parkinson’s disease – Cognitive Functional Rating Scale; ToL = Tower of London.

### 13 Exploratory univariate linear mixed-model analyses of neuropsychological test outcomes at six-months follow-up

**Table S14:** Group differences at follow-up (T2) from the exploratory univariate linear mixed-model analyses on the individual neuropsychological test outcomes for the crude and adjusted analysis models in the intention-to-treat sample.

| | T2 | | Group difference (crude model) | | | | Group difference (adjusted model)* | | | |
|---|---|---|---|---|---|---|---|---|---|---|
| | Active control M (SD) | Cognitive training M (SD) | B [SE] | 95% CI | *P* value (FDR) | *P* value (raw) | B [SE] | 95% CI | *P* value (FDR) | *P* value (raw) |
| ToL S1 accuracy (%, N=126) | 96.3 (4.5) | 96.1 (7.1) | 1.11 [0.83] | -0.52 to 2.74 | 0.816 | 0.183 | 1.13 [0.82] | -0.47 to 2.74 | 0.820 | 0.166 |
| ToL S2 accuracy (%, N=126) | 92.2 (8.7) | 90.1 (17.3) | 1.29 [1.67] | -1.99 to 4.58 | 0.816 | 0.438 | 1.34 [1.66] | -1.93 to 4.61 | 0.820 | 0.419 |
| ToL S3 accuracy (%, N=126) | 88.4 (11.2) | 86.0 (17.7) | 1.09 [1.84] | -2.53 to 4.72 | 0.816 | 0.553 | 0.59 [1.85] | -3.05 to 4.23 | 0.893 | 0.750 |
| ToL S4 accuracy (%, N=126) | 77.1 (16) | 74.0 (22.5) | 0.94 [2.68] | -4.33 to 6.21 | 0.873 | 0.727 | 0.55 [2.66] | -4.69 to 5.79 | 0.893 | 0.835 |
| ToL S5 accuracy (%, N=126) | 67.3 (20.8) | 60.1 (25.1) | -2.68 [2.94] | -8.48 to 3.11 | 0.816 | 0.363 | -3.57 [2.91] | -9.31 to 2.16 | 0.820 | 0.221 |
| ToL S1 reaction time (s, N=126) | 5.9 (2.1) | 5.8 (2.9) | -0.08 [0.25] | -0.57 to 0.40 | 0.873 | 0.732 | 0.03 [0.24] | -0.44 to 0.51 | 0.925 | 0.895 |
| ToL S2 reaction time (s, N=126) | 7.7 (2.7) | 7.4 (3.2) | -0.25 [0.27] | -0.78 to 0.29 | 0.816 | 0.364 | -0.20 [0.27] | -0.73 to 0.32 | 0.820 | 0.446 |
| ToL S3 reaction time (s, N=126) | 10.3 (3.4) | 10.1 (4.1) | -0.40 [0.37] | -1.13 to 0.33 | 0.816 | 0.282 | -0.28 [0.37] | -1.01 to 0.45 | 0.820 | 0.457 |
| ToL S4 reaction time (s, N=126) | 14.3 (4.6) | 14.8 (5.7) | 0.10 [0.58] | -1.04 to 1.23 | 0.918 | 0.868 | 0.33 [0.57] | -0.80 to 1.45 | 0.820 | 0.568 |
| ToL S5 reaction time (s, N=125) | 20.4 (4.9) | 21.0 (5.4) | 0.58 [0.75] | -0.89 to 2.05 | 0.816 | 0.438 | 0.75 [0.74] | -0.71 to 2.21 | 0.820 | 0.311 |
| MoCA (N=132) | 26.8 (2.3) | 27.3 (2.2) | 0.31 [0.36] | -0.40 to 1.01 | 0.816 | 0.396 | 0.20 [0.35] | -0.48 to 0.89 | 0.820 | 0.560 |
| SCWT card I (s, N=132) | 52.7 (11.5) | 51.0 (11.7) | 0.23 [1.37] | -2.47 to 2.92 | 0.918 | 0.869 | 0.77 [1.34] | -1.87 to 3.41 | 0.820 | 0.566 |
| SCWT card II (s, N=132) | 66.1 (14) | 64.6 (15) | -1.25 [1.66] | -4.52 to 2.02 | 0.816 | 0.452 | -0.92 [1.63] | -4.14 to 2.30 | 0.820 | 0.574 |
| SCWT card III (s, N=132) | 102.6 (26.1) | 102.2 (27.8) | -4.24 [3.25] | -10.65 to 2.17 | 0.816 | 0.194 | -3.28 [3.22] | -9.62 to 3.06 | 0.820 | 0.309 |
| SCWT interference score (N=132)[#] | 1.6 (0.2) | 1.6 (0.3) | -0.01 [0.04] | -0.08 to 0.07 | 0.918 | 0.888 | -0.01 [0.04] | -0.08 to 0.06 | 0.893 | 0.829 |
| Letter fluency (N=132) | 44.1 (13.2) | 42.5 (12.3) | -1.42 [1.29] | -3.97 to 1.13 | 0.816 | 0.274 | -1.69 [1.29] | -4.23 to 0.85 | 0.820 | 0.191 |
| Category fluency (N=132) | 22.9 (6.2) | 21.6 (5.1) | -0.51 [0.76] | -2.01 to 1.00 | 0.816 | 0.507 | -0.66 [0.75] | -2.14 to 0.83 | 0.820 | 0.384 |
| RCFT (N=132) | 30.9 (4.1) | 31.0 (3.6) | -0.22 [0.50] | -1.21 to 0.76 | 0.845 | 0.654 | -0.29 [0.50] | -1.28 to 0.70 | 0.820 | 0.562 |
| RAVLT learning curve (N=132)[†] | 0.3 (0.1) | 0.3 (0.2) | -0.02 [0.02] | -0.06 to 0.03 | 0.816 | 0.532 | -0.02 [0.02] | -0.07 to 0.03 | 0.820 | 0.417 |
| RAVLT direct recall (N=132) | 45.1 (10.7) | 45.4 (11.8) | 0.62 [1.33] | -2.00 to 3.24 | 0.845 | 0.644 | 0.35 [1.32] | -2.24 to 2.95 | 0.893 | 0.789 |
| RAVLT delayed recall (N=132) | 9.7 (3.7) | 9.8 (3.4) | 0.09 [0.43] | -0.76 to 0.94 | 0.918 | 0.835 | 0.00 [0.42] | -0.83 to 0.83 | 1.000 | 1.000 |
| RAVLT recognition (N=132) | 28.8 (1.7) | 28.6 (2.2) | -0.25 [0.31] | -0.86 to 0.36 | 0.816 | 0.424 | -0.25 [0.31] | -0.85 to 0.36 | 0.820 | 0.424 |
| Digit span forward score (N=132) | 9.4 (1.8) | 9.3 (1.8) | -0.02 [0.24] | -0.50 to 0.46 | 0.932 | 0.932 | -0.12 [0.24] | -0.58 to 0.35 | 0.846 | 0.629 |
| Digit span forward span length (N=132) | 6.3 (1.2) | 6.2 (1.1) | -0.11 [0.15] | -0.42 to 0.19 | 0.816 | 0.472 | -0.17 [0.15] | -0.47 to 0.12 | 0.820 | 0.252 |
| Digit span backward score (N=132) | 7.0 (1.8) | 7.1 (1.2) | 0.35 [0.25] | -0.13 to 0.84 | 0.816 | 0.155 | 0.36 [0.24] | -0.12 to 0.84 | 0.820 | 0.143 |
| Digit span backward span length (N=132) | 5.0 (1.1) | 5.0 (0.8) | 0.07 [0.16] | -0.24 to 0.38 | 0.845 | 0.649 | 0.06 [0.16] | -0.25 to 0.36 | 0.874 | 0.705 |
| LLT learning curve (N=131) | 0.7 (0.3) | 0.7 (0.3) | -0.04 [0.04] | -0.13 to 0.04 | 0.816 | 0.324 | -0.04 [0.04] | -0.12 to 0.04 | 0.820 | 0.311 |
| LLT direct recall (N=131) | 17.4 (17.6) | 18.8 (20.8) | 1.84 [2.75] | -3.58 to 7.25 | 0.816 | 0.505 | 1.48 [2.68] | -3.81 to 6.76 | 0.820 | 0.582 |
| LLT delayed recall (N=130) | 0.8 (1.7) | 1.0 (2.7) | 0.27 [0.43] | -0.59 to 1.13 | 0.816 | 0.535 | 0.19 [0.43] | -0.65 to 1.03 | 0.846 | 0.655 |
| BNT (N=132) | 57.3 (2.4) | 56.5 (2.9) | -0.57 [0.32] | -1.20 to 0.07 | 0.816 | 0.081 | -0.55 [0.33] | -1.19 to 0.09 | 0.820 | 0.092 |
| BVFDT (N=132) | 29.5 (2.5) | 30.4 (2.2) | 0.78 [0.40] | -0.01 to 1.56 | 0.816 | 0.054 | 0.81 [0.40] | 0.03 to 1.59 | 0.820 | **0.042** |

Significant differences are marked in bold. *Corrected for age, sex and education in years; [#]Stroop interference score calculation: Stroop card III time / Stroop card II time;[30] [†]RAVLT learning curve calculated similar to the LLT learning curve model.[9] *Abbreviations:* BNT – Boston Naming Test; BVFDT – Benton Visual Form Discrimination Test; MoCA – Montreal Cognitive Assessment; RAVLT – Rey Auditory Verbal Learning Test; RCFT – Rey Complex Figure Test; SCWT – Stroop Color Word Test; S1-S5 – ToL difficulty load 1-5; ToL – Tower of London.

**14 Post-hoc analyses of differential CT effects in PD patients with normal cognition, mild cognitive impairment, or dementia**

The differential effects of CT relative to AC in PD patients with normal cognition (PD-NC), mild cognitive impairment (PD-MCI) or dementia (PD-D) are illustrated in Figure S3 and statistics are reported in Table S16 and Table S17.

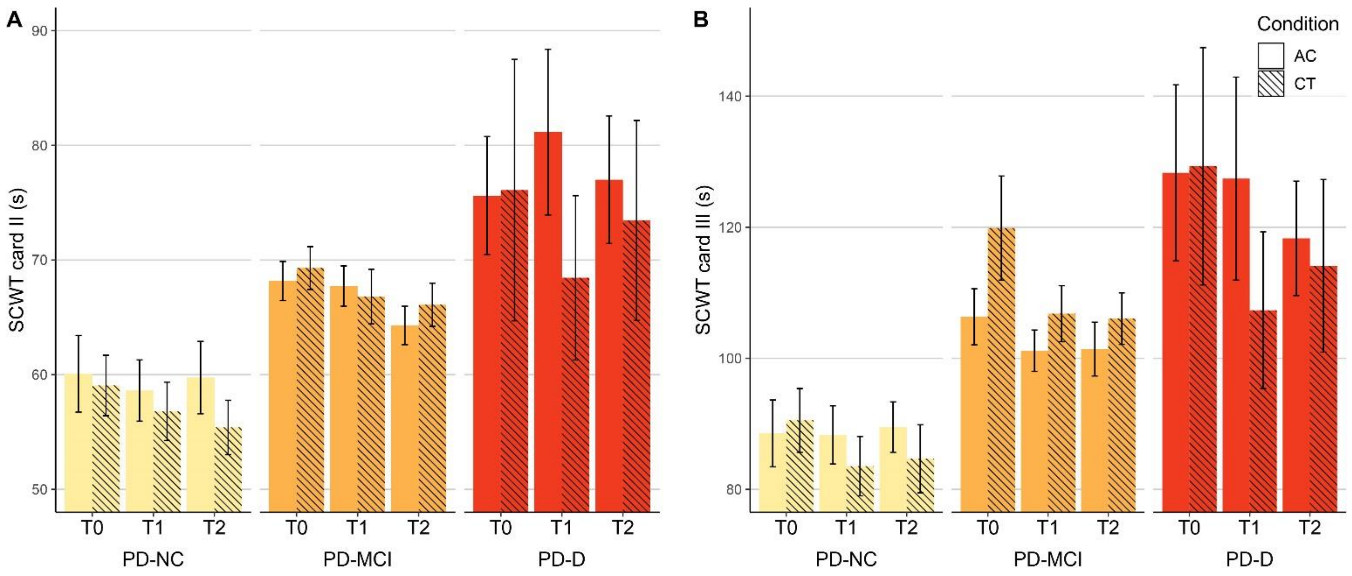


**Figure S3:** Difference between the CT and AC group on the Stroop color word test card II **(A)** and III **(B)**, separated for participants with normal cognition (PD-NC), PD-mild cognitive impairment (PD-MCI) and PD dementia (PD-D). Data shown are observed means ± standard error.

**Table S15:** Post-hoc analyses of group differences after training (T1) for participants with PD normal cognition, PD-MCI and PD-D from the multivariate linear mixed-model analyses on the primary outcome measure and outcome measures that responded to CT.
The top part shows group differences (CT versus active control) per sub-group of participants. Positive estimates for ToL accuracy and negative estimates for ToL reaction time and the SCWT indicate effects in favor of CT. The lower part shows interaction effects, i.e. the difference between the three sub-groups on the CT effect. Positive estimates for ToL accuracy and negative estimates for ToL reaction time and the SCWT indicate added positive effects of CT for the specified contrast.

| | | **MAIN EFFECTS at T1** | | | | | | | | |
|---|---|---|---|---|---|---|---|---|---|---|
| | | **PD-NC** | | | **PD-MCI** | | | **PD-D** | | |
| | | B [SE] | 95% CI | p-value | B [SE] | 95% CI | p-value | B [SE] | 95% CI | p-value |
| Overall ToL accuracy | | -0.206 [0.210] | -0.622 to 0.210 | 0.329 | -0.153 [0.123] | -0.397 to 0.092 | 0.219 | **0.582 [0.281]** | **0.025 to 1.139** | **0.041** |
| Sub-score: | S1 | -0.104 [0.336] | -0.763 to 0.555 | 0.757 | -0.161 [0.197] | -0.548 to 0.225 | 0.413 | 0.272 [0.449] | -0.610 to 1.154 | 0.545 |
| | S2 | -0.302 [0.335] | -0.960 to 0.357 | 0.369 | -0.200 [0.197] | -0.586 to 0.187 | 0.310 | 0.310 [0.449] | -0.572 to 1.193 | 0.490 |
| | S3 | -0.125 [0.335] | -0.784 to 0.534 | 0.709 | -0.016 [0.197] | -0.402 to 0.370 | 0.936 | 0.454 [0.449] | -0.428 to 1.336 | 0.312 |
| | S4 | -0.366 [0.335] | -1.025 to 0.293 | 0.276 | -0.251 [0.197] | -0.638 to 0.135 | 0.202 | **0.897 [0.449]** | **0.014 to 1.779** | **0.046** |
| | S5 | -0.112 [0.335] | -0.770 to 0.546 | 0.738 | -0.162 [0.197] | -0.549 to 0.225 | 0.410 | **0.988 [0.449]** | **0.106 to 1.870** | **0.028** |
| Overall ToL reaction time | | -0.195 [0.204] | -0.599 to 0.209 | 0.342 | -0.094 [0.120] | -0.331 to 0.143 | 0.434 | -0.373 [0.273] | -0.915 to 0.169 | 0.175 |
| Sub-score: | S1 | 0.033 [0.259] | -0.478 to 0.544 | 0.899 | -0.131 [0.152] | -0.430 to 0.168 | 0.389 | **-0.689 [0.348]** | **-1.374 to -0.005** | **0.049** |
| | S2 | -0.143 [0.259] | -0.654 to 0.367 | 0.581 | -0.064 [0.152] | -0.364 to 0.236 | 0.674 | -0.154 [0.349] | -0.841 to 0.532 | 0.658 |
| | S3 | -0.240 [0.259] | -0.751 to 0.270 | 0.355 | -0.018 [0.152] | -0.317 to 0.282 | 0.908 | -0.566 [0.347] | -1.250 to 0.117 | 0.104 |
| | S4 | -0.216 [0.260] | -0.727 to 0.295 | 0.406 | -0.246 [0.152] | -0.546 to 0.054 | 0.108 | **-0.694 [0.347]** | **-1.378 to -0.011** | **0.047** |
| | S5 | -0.422 [0.259] | -0.933 to 0.088 | 0.105 | -0.003 [0.153] | -0.303 to 0.297 | 0.984 | 0.209 [0.348] | -0.476 to 0.894 | 0.549 |
| SCWT card I | | -1·048 [2·849] | -6·683 to 4·588 | 0·714 | -1·406 [1·653] | -4·676 to 1·864 | 0·396 | -5·383 [3·265] | -11·841 to 1·075 | 0·102 |
| SCWT card II | | -1·086 [3·849] | -8·699 to 6·528 | 0·778 | -1·834 [2·232] | -6·249 to 2·582 | 0·413 | **-14·432 [4·407]** | **-23·148 to -5·716** | **0·001** |
| SCWT card III | | -5·778 [6·968] | -19·562 to 8·006 | 0·409 | -1·296 [4·077] | -9·361 to 6·769 | 0·751 | **-21·076 [8·109]** | **-37·116 to -5·036** | **0·010** |
| | | **INTERACTION EFFECTS at T1** | | | | | | | | |
| | | **PD-MCI > PD-NC** | | | **PD-D > PD-NC** | | | **PD-D > PD-MCI** | | |
| | | B [SE] | 95% CI | p-value | B [SE] | 95% CI | p-value | B [SE] | 95% CI | p-value |
| Overall ToL accuracy | | 0.053 [0.244] | -0.430 to 0.536 | 0.828 | **0.788 [0.351]** | **0.093 to 1.482** | **0.027** | **0.734 [0.307]** | **0.126 to 1.343** | **0.018** |
| Overall ToL reaction time | | 0.101 [0.237] | -0.368 to 0.570 | 0.670 | -0.178 [0.340] | -0.853 to 0.496 | 0.601 | -0.279 [0.299] | -0.871 to 0.313 | 0.352 |
| SCWT I | | -0·359 [3·294] | -6·874 to 6·156 | 0·913 | -4·335 [4·333] | -12·906 to 4·235 | 0·319 | -3·976 [3·656] | -11·208 to 3·256 | 0·279 |
| SCWT II | | -0·748 [4·451] | -9·552 to 8·055 | 0·867 | **-13·346 [5·852]** | **-24·922 to -1·771** | **0·024** | **-12·598 [4·938]** | **-22·365 to -2·832** | **0·012** |
| SCWT III | | 4·482 [8·068] | -11·477 to 20·442 | 0·579 | -15·298 [10·691] | -36·445 to 5·850 | 0·155 | **-19·780 [9·071]** | **-37·724 to -1·836** | **0·031** |

Significant differences are marked in bold. *Abbreviations:* PD-D – Parkinson’s disease – dementia; PD-MCI – Parkinson’s disease – mild cognitive impairment; PD-NC – Parkinson’s disease – normal cognition; SCWT – Stroop Color Word Test; S1-S5 – ToL difficulty load 1-5; ToL – Tower of London.

**Table S16:** Post-hoc analyses of group differences after training (T2) for participants with PD normal cognition, PD-MCI and PD-D from the multivariate linear mixed-model analyses on the primary outcome measure and outcome measures that responded to CT.
The top part shows group differences (CT versus active control) per sub-group of participants. Positive estimates for ToL accuracy and negative estimates for ToL reaction time and the SCWT indicate effects in favor of CT. The lower part shows interaction effects, i.e. the difference between the three sub-groups on the CT effect. Positive estimates for ToL accuracy and negative estimates for ToL reaction time and the SCWT indicate added positive effects of CT for the specified contrast.

| | | **MAIN EFFECTS - CRUDE MODEL - T2** | | | | | | | | |
|---|---|---|---|---|---|---|---|---|---|---|
| | | **PD-NC** | | | **PD-MCI** | | | **PD-D** | | |
| | | B [SE] | 95% CI | p-value | B [SE] | 95% CI | p-value | B [SE] | 95% CI | p-value |
| Overall ToL accuracy | | -0.092 [0.197] | -0.481 to 0.296 | 0.639 | -0.104 [0.122] | -0.344 to 0.136 | 0.392 | **0.804 [0.266]** | **0.279 to 1.329** | **0.003** |
| Sub-score: | S1 | 0.333 [0.332] | -0.319 to 0.986 | 0.316 | -0.140 [0.200] | -0.533 to 0.253 | 0.484 | **1.327 [0.437]** | **0.470 to 2.185** | **0.002** |
| | S2 | 0.120 [0.332] | -0.532 to 0.772 | 0.718 | -0.199 [0.200] | -0.592 to 0.194 | 0.321 | **1.281 [0.437]** | **0.423 to 2.139** | **0.003** |
| | S3 | -0.171 [0.332] | -0.823 to 0.482 | 0.608 | -0.092 [0.200] | -0.485 to 0.300 | 0.645 | **0.872 [0.437]** | **0.014 to 1.730** | **0.046** |
| | S4 | -0.143 [0.332] | -0.795 to 0.510 | 0.668 | -0.168 [0.200] | -0.561 to 0.225 | 0.401 | **0.917 [0.437]** | **0.059 to 1.775** | **0.036** |
| | S5 | -0.574 [0.332] | -1.227 to 0.078 | 0.084 | -0.230 [0.200] | -0.623 to 0.163 | 0.252 | 0.541 [0.437] | -0.317 to 1.399 | 0.216 |
| Overall ToL reaction time | | -0.053 [0.196] | -0.442 to 0.335 | 0.787 | 0.096 [0.118] | -0.137 to 0.329 | 0.417 | -0.213 [0.264] | -0.736 to 0.309 | 0.420 |
| Sub-score: | S1 | 0.001 [0.259] | -0.509 to 0.511 | 0.997 | 0.046 [0.155] | -0.259 to 0.350 | 0.769 | -0.505 [0.343] | -1.179 to 0.170 | 0.142 |
| | S2 | -0.098 [0.259] | -0.607 to 0.412 | 0.706 | 0.012 [0.155] | -0.293 to 0.316 | 0.940 | -0.323 [0.343] | -0.998 to 0.353 | 0.348 |
| | S3 | 0.008 [0.259] | -0.501 to 0.518 | 0.974 | -0.013 [0.155] | -0.317 to 0.291 | 0.935 | -0.336 [0.343] | -1.010 to 0.338 | 0.328 |
| | S4 | 0.161 [0.259] | -0.349 to 0.671 | 0.534 | 0.062 [0.155] | -0.242 to 0.367 | 0.688 | -0.101 [0.343] | -0.775 to 0.574 | 0.769 |
| | S5 | 0.031 [0.259] | -0.479 to 0.541 | 0.905 | 0.281 [0.155] | -0.025 to 0.586 | 0.071 | 0.120 [0.343] | -0.555 to 0.794 | 0.727 |
| SCWT card I | | -1·940 [2·893] | -7·638 to 3·758 | 0·503 | 0·843 [1·705] | -2·516 to 4·201 | 0·622 | 1·357 [3·242] | -5·026 to 7·740 | 0·676 |
| SCWT card II | | -2·411 [3·475] | -9·254 to 4·432 | 0·488 | 0·855 [2·046] | -3·175 to 4·885 | 0·676 | -5·279 [3·890] | -12·939 to 2·382 | 0·176 |
| SCWT card III | | -4·608 [6·858] | -18·114 to 8·898 | 0·502 | -2·482 [4·060] | -10·476 to 5·513 | 0·542 | -5·765 [7·679] | -20·887 to 9·357 | 0·454 |
| | | **INTERACTION EFFECTS - CRUDE MODEL - T2** | | | | | | | | |
| | | **PD-MCI > PD-NC** | | | **PD-D > PD-NC** | | | **PD-D > PD-MCI** | | |
| | | B [SE] | 95% CI | p-value | B [SE] | 95% CI | p-value | B [SE] | 95% CI | p-value |
| Overall ToL accuracy | | -0.012 [0.227] | -0.460 to 0.436 | 0.958 | **0.897 [0.328]** | **0.249 to 1.544** | **0.007** | **0.909 [0.289]** | **0.338 to 1.479** | **0.002** |
| Overall ToL reaction time | | 0.149 [0.227] | -0.300 to 0.598 | 0.513 | -0.160 [0.327] | -0.808 to 0.488 | 0.625 | -0.309 [0.288] | -0.879 to 0.260 | 0.285 |
| SCWT I | | 2·782 [3·358] | -3·829 to 9·394 | 0·408 | 3·297 [4·344] | -5·257 to 11·851 | 0·449 | 0·514 [3·660] | -6·693 to 7·722 | 0·888 |
| SCWT II | | 3·266 [4·034] | -4·678 to 11·210 | 0·419 | -2·868 [5·218] | -13·143 to 7·408 | 0·583 | -6·134 [4·395] | -14·788 to 2·520 | 0·164 |
| SCWT III | | 2·126 [7·971] | -13·570 to 17·822 | 0·790 | -1·157 [10·296] | -21·432 to 19·119 | 0·911 | -3·283 [8·682] | -20·380 to 13·814 | 0·706 |

Significant differences are marked in bold. *Abbreviations:* PD-D – Parkinson's disease – dementia; PD-MCI – Parkinson's disease – mild cognitive impairment; PD-NC – Parkinson's disease – normal cognition; SCWT – Stroop Color Word Test; S1-S5 – ToL difficulty load 1-5; ToL – Tower of London.
